# Psychedelics as pharmacotherapeutics for substance use disorders: a scoping review on clinical trials and perspectives on underlying neurobiology

**DOI:** 10.1101/2025.04.04.25324315

**Authors:** Lucas Wittenkeller, Gary Gudelsky, John T. Winhusen, Davide Amato

**Author notes:** Corresponding Author Contact Information: Dr. Davide Amato, Address: 231 Albert Sabin Way, Cincinnati, Ohio, 45267.

## Abstract

Psychedelics have garnered great attention in recent years as treatments for major depressive disorder and treatment-resistant depression due to their ability to alter consciousness and afflicted cognitive processes with lasting effects. Given these unique characteristics and the urgent need for efficacious treatments, psychedelics are being tested for a variety of psychiatric conditions, including substance use disorders (SUDs). Despite promising results and growing interest, the neurobiological mechanisms underlying therapeutic efficacy of psychedelics remain uncharacterized. Using a scoping review approach, we summarize current clinical trials registered at ClinicalTrials.gov that utilize classic psychedelics as interventions for SUDs with the goal of understanding the current state and outlook of the field. A second scoping review was conducted using PubMed and SCOPUS databases to identify the relevant publications addressing the pharmacotherapeutic potential of restoring dopamine homeostasis as a novel neurobiological mechanism of psychedelics. This mechanism may blunt drug-seeking behavior, promote drug abstinence, and underlie their clinical relevance for SUD in addition to previously characterized mechanisms.

## Introduction

Substance use disorder (SUD) is a complex condition characterized by the uncontrollable urge to use drugs despite negative consequences and is a major cause of death and disability worldwide. In the United States in 2020, the one-year prevalence of SUDs was estimated at around 17.7% in individuals with no other psychiatric conditions (1) resulting in an economic burden of up to $35.3 billion annually (2). Pharmacotherapeutics are not available for all SUDs. Specifically, there are thirteen FDA-approved medications that treat opioid, alcohol, and/or nicotine use disorder (3), while medications for cannabis, cocaine, methamphetamine, or inhalant use disorders are still lacking (4). The supervised use of psychedelic drugs in the context of psychotherapy, known as psychedelic-assisted psychotherapy, is currently garnering attention in the field for its therapeutic promise in managing symptoms of psychiatric illnesses including SUDs (5–8). In the present review, we describe ongoing clinical trials investigating the effectiveness of psychedelic drugs in treating symptoms associated with the relapsing use of alcohol, nicotine, opioids, methamphetamine, cannabis, and cocaine. We also discuss potential mechanisms underlying the therapeutic efficacy of psychedelics in substance use disorders (SUDs), with a particular emphasis on the restoration of dopamine homeostasis as a novel pharmacotherapeutic mechanism that may reduce drug-seeking and drug-taking behaviors.

To fully address these objectives, two scoping reviews were conducted: one to capture all registered clinical trials utilizing psychedelics to treat SUDs, and another to identify publications reporting psychedelic-induced dopaminergic changes in the nucleus accumbens via *in vivo* microdialysis in animals, with the aim of characterizing dopaminergic effects across this class of drugs. Both scoping reviews were conducted in accordance with PRISMA-ScR guidelines (9). All methodological details supporting Table 1 and Figure 3 are provided in the supplementary documentation.

**Table 1:**
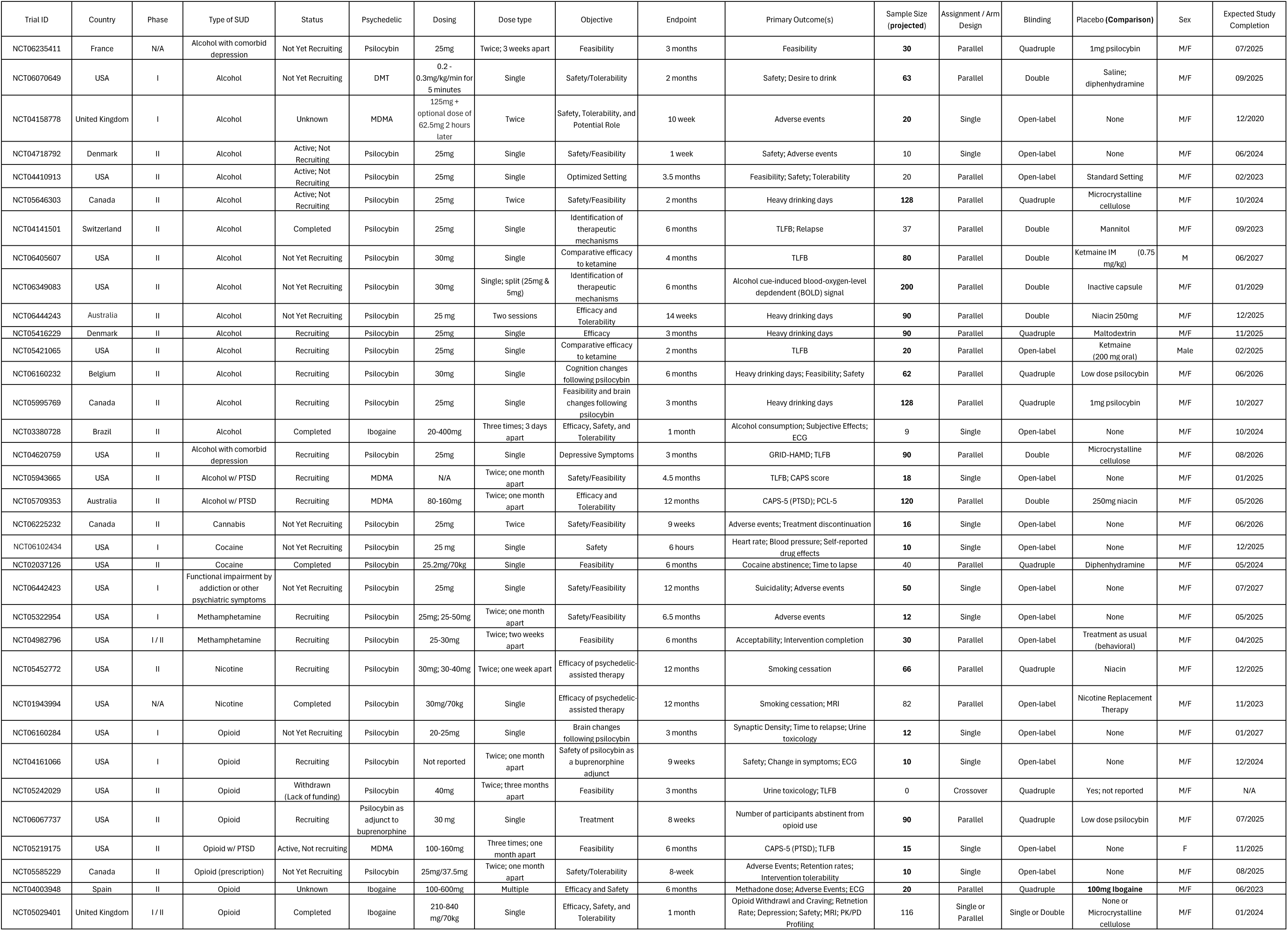
Registered Clinical Trials Investigating Psychedelic-Assisted Treatment for SUDs.

### Current State of Clinical Research on Psychedelic-Assisted Interventions

The term ‘psychedelics’ typically refers to classic compounds including psilocybin, lysergic acid diethylamide (LSD), *N*,*N*-dimethyltryptamine (DMT), and mescaline, which are agonists of the serotonin 2A (5-HT_2A_) receptor. Psychedelics were first explored for efficacy in treating SUDs beginning in the 1950s and showed therapeutic promise in both narcotic (10) and alcohol (11) use disorders. By 1970, about 40,000 individuals received psychedelic treatment resulting in more than 1000 publications (12). Despite this, research using psychedelics came to a halt following the passage of the Controlled Substances Act in 1971, which placed psychedelics under Schedule I classification indicating their lack of therapeutic benefit and high probability of abuse. Since 2000, in response to studies demonstrating lack of toxicity (13, 14) or abuse potential (15) of psychedelic compounds, researchers have witnessed a resurgence of interest in the potential for psychedelics to ameliorate steadily increasing rates of drug and alcohol misuse. Importantly, when we use the term ‘psychedelic’, we are referring strictly to the classic psychedelics as this is not always the case for some hallucinogens or dissociative anesthetics, like ketamine, that is often discussed with psychedelics and has been shown to be efficacious for SUDs (16) despite having abuse liability itself (17) Therefore, the safety and lack of abuse potential pose classic psychedelics as a desirable treatment option for SUDs and other psychiatric illnesses.

The psychedelic that has earned the most attention as a pharmacotherapeutic for psychiatric conditions is psilocybin, the psychoactive component of “magic mushrooms” (18). Psilocybin-assisted psychotherapy has been awarded Breakthrough Therapy status by the FDA for the treatment of major depressive disorder (MDD) (19) where clinical trials have found decreases in depression (20–22) lasting up to one year (23). Since then, psilocybin-assisted psychotherapy has been tested in clinical studies as a potential treatment intervention for SUDs and other psychopathologies (8). Two foundational open-label studies have highlighted the potential for psychedelic interventions to be used for SUDs (24, 25). Johnson and colleagues demonstrated that administration of 20mg/70kg, 30mg/70kg, and 30mg/70kg psilocybin at weeks 5, 7, and 13 of a treatment plan for smoking cessation, respectively, decreased daily mean cigarette consumption six months after psychedelic administration, with 11 of 15 participants reporting smoking cessation after the first psilocybin session and all other participants reporting decreased cigarettes per day compared to pre-psilocybin levels (25). At 12- and 30-months follow-up, 10 and 9 individuals, respectively, maintained drug abstinence (26). Similarly, Bogenschutz and colleagues showed that 21mg/70kg and 28mg/70kg of psilocybin decreased percentage of drinking and heavy drinking days for up to 32 weeks compared to baseline (24). Because this study was investigating feasibility of psilocybin-assisted treatment for alcohol use disorder (AUD), it lacked a control group and utilized a small sample size of ten individuals. To address this, Bogenschutz and colleagues conducted a double-blind, randomized, controlled trial (RCT) evaluating the effect of two sessions of psychedelic-assisted manualized psychotherapy where psilocybin was administered (25mg/70kg and 25-40mg/70kg) to 95 individuals with AUD. The results revealed that psilocybin, relative to diphenhydramine control, decreased percentage of drinking days and number of drinks per day during the 32-week study period (27).

In 2022, following publication of the first evidence outlining the lasting effectiveness of psychedelic intervention for decreasing alcohol and nicotine consumption, there were 11 active RCTs evaluating psychedelic-assisted treatment for alcohol, nicotine, opioid, or methamphetamine use disorders (28). Due to the novelty of applying psychedelics to SUDs, many clinical trials employ experimental designs that have been excluded from prior systematic literature reviews, including open-label or single assignment designs. Therefore, we sought to include and describe all clinical trials currently registered to better understand the current outlook of the field. Below we describe the research design, primary and secondary outcomes, and purpose for each ongoing clinical trial. We identified registered clinical trials on ClinicalTrials.gov using condition criteria of substance use disorder, addiction, and substance-related disorders and intervention criteria of psychedelic, psilocybin, DMT, LSD, mescaline, and MDMA in August 2024. Clinical trials were excluded if repeated, lacking a psychedelic intervention, or not including individuals with substance use disorder(s). Clinical trials applying MDMA or ibogaine, a hallucinogen and non-classic psychedelic respectively, were included for their shared ability to bind serotonin receptors (29, 30) and similar application for treating psychiatric disorders (30, 31) despite not being a classic psychedelic compound in the strictest sense. Other classic psychedelics were included in our search, however only one additional trial was identified (NCT06070649). Because these clinical trials are ongoing with results not posted yet, it is not possible to extensively discuss each subtype of SUD. Thus, we have organized Table 1 by its subclass of SUD but will discuss all clinical trials together for simplicity.

### Application and Experimental Design

Thirty-four clinical trials were identified consisting of two pilot studies, seven phase I studies, two phase I/II study, and 23 phase II studies (**Table 1**). Of these studies, 18 focused on AUD, eight on opioid use disorder (OUD), two on nicotine use disorder, two on methamphetamine use disorder, two on cocaine use disorder, and one on cannabis use disorder. Four of the 17 AUD trials are designed to evaluate psychedelic use for AUD and a co-occurring condition (depression or post-traumatic stress disorder, PTSD). One OUD trial will evaluate MDMA for comorbid OUD and PTSD. One study will not measure a particular SUD, but rather functional impairment caused by mood and anxiety disorders, trauma, or SUD. MDMA intervention studies include three of the 17 studies on AUD and one of six studies on OUD. The heavy focus on AUD in clinical trials may be due to the high comorbidity of major depressive disorder (MDD) with AUD (32, 33) and psilocybin’s recent success in treating MDD (19). Ibogaine intervention makes up three of 34 clinical trials and are focused on OUD or AUD. The lack of clinical trials utilizing ibogaine is likely due to concerns for participant safety (30). Despite psilocybin’s success in clinical trials, it has been suggested that clinical trials utilize other classical psychedelics that share mechanisms of action, but with a shorter duration of action, such as LSD, DMT, or mescaline (34). To this end, one of 17 clinical trials testing psychedelic efficacy in AUD is utilizing DMT.

Although RCTs are the gold standard for clinical trials based on their ability to evaluate causal relationships by eliminating biases inherent in many other designs (35), it has been suggested that recent emphasis on one type of experimental design ignores the importance and necessity of other approaches to more appropriately answer particular research questions (36). Of the 34 registered trials, 17 will evaluate the efficacy of psychedelic treatment relative to a non-treatment control (placebo) on substance use as a primary outcome. Of these 17 trials, 15 utilize parallel assignment and are blinded and placebo controlled. An area lacking consensus among these RCTs is the choice of placebo, as there are six different placebos used across these studies including low-dose psilocybin, mannitol, niacin, microcrystalline cellulose, diphenhydramine, and maltodextrin. Three trials have the goal of comparing the efficacy of a psychedelic to an active comparator with two trials comparing psilocybin to ketamine for AUD and one comparing psilocybin to nicotine replacement therapy for smoking-cessation. Fifteen trials have a primary objective of testing the feasibility, safety, and/or tolerability of psychedelic treatment with 9 trials using a single arm open-label design. Two trials evaluating a psychedelic for use in SUD and PTSD have PTSD as the primary outcome measure (NCT05709353; NCT05219175). One trial is designed to compare two “set and setting” protocols in combination with psilocybin for the treatment of AUD (NCT04410913). Two trials are evaluating the use of a psychedelic with another medication, with one evaluating psilocybin as an adjunct to buprenorphine for the treatment of OUD (NCT06067737) and one evaluating the ability of psilocybin to help individuals taper from long-term opioid therapy for chronic pain (NCT05585229). The trial with the largest target sample size (N=200) is a mechanistic clinical trial of psilocybin for AUD with a focus on evaluating the potential neural mechanisms by which psilocybin may improve AUD outcomes (NCT06349083).

Treatment interventions being evaluated include psilocybin (26/34), MDMA (4/34), ibogaine (3/34), and DMT (1/34). Doses under examination are 20-40 mg for psilocybin, 0.2-0.3 mg/kg for DMT, and 80-160 mg for MDMA. For clinical trials identified in Table 1, psilocybin is administered once, repeated, or escalating; DMT is administered once, and MDMA is administered repeatedly. Psilocybin and DMT doses tested for treatment of SUDs reflect therapeutic dose thresholds previously established for MDD (21, 37, 38). Similarly, standard MDMA-assisted psychotherapy for PTSD involves an initial 125 mg dose followed by an offer of an additional half dose two hours after administration (31), which falls within the 80-160 mg dose range used for MDMA-assisted treatment of SUDs. Most studies have early study status with 10 not yet recruiting, 12 recruiting, four active and not recruiting, five completed, two with unknown status, and one withdrawn. Of the five completed clinical trials, none have posted results and only one trial has submitted results.

### Outcomes

Experimental outcomes of clinical trials evaluating psychedelics as treatment for SUDs mostly involve measures of drug use gathered using Timeline Follow Back assessments (TLFB); comorbid symptomology and subjective effects measured using behavioral scales, and neurobiological changes measured using functional magnetic resonance imaging (fMRI) or positron emission tomography (PET) imaging. Because of the phase I and II statuses for these ongoing trials, many studies aim to measure safety, tolerability, feasibility, and compliance in addition to changes in drug use across subpopulations of individuals suffering from different SUDs. Common outcomes to be assessed include adverse events, electrocardiogram, participant retention, and treatment compliance. One study testing feasibility and safety of psilocybin is also developing a pharmacokinetic profile for psilocin, the active metabolite of psilocybin, for further safety and efficacy analyses (NCT04718792). These measures inform researchers on dose tolerability and safety of treatment regimens, validity of self-reported scales of drug use, and likelihood of treatment adherence.

Treatment-related effects, not including adverse events, are measurements of subjective experiences induced by psychedelic administration that are collected using self-rated questionnaires, such as the Mystical Experience Questionnaire (MEQ) -30 and -43 that are derived from the States of Consciousness Questionnaire (39). MEQ is most used for its dose-dependent reactivity (40) and prediction of therapeutic outcomes (24). These outcomes measure constructs relating to mystical experiences, spirituality, mood, perception, and unity. Since it is impossible to fully measure and understand the subjective experience of psychedelics, this is currently the field’s best standardized metric for participant experience and outlook.

Drug-related behavioral outcomes measure treatment-onset changes by comparing various follow-ups to baseline responses. These behaviors can include measures of time to first drug use, amount and frequency of drug use, craving, cessation, and abstinence. TLFB is a popular calendar-based, self-reported measure originally designed for AUD where data are reported as percentage of days drinking and number of days of heavy drinking (41). The use of TLFB has high validity for tobacco, cannabis, and cocaine use and has been proposed to limit the need for biological samples (41, 42). Therefore, it has been widely adopted in clinical trials for its validity and low cost and energy demands by participants and research staff. Nonetheless, biological samples, such as measures of carbon monoxide for recent cigarette smoking or breath alcohol levels for recent drinking, urine drug screens for illicit substance use, or PET scans of drug-afflicted brain regions (43) are still used in combination with self-reports, which are considered gold standard in identifying and assessing SUDs.

Depression and trauma-related behavior scales are sometimes administered in trials measuring SUD behavioral patterns likely due to high incidence of comorbidity of SUDs with other psychiatric illnesses (32, 33). Therefore, it is not surprising that these clinical trials utilize similar behavioral scales used in MDD and PTSD clinical trials like Beck’s Depression Inventory (BDI), Quick Inventory of Depressive Symptomatology (QIDS-SR), and GRID-HAM-D for depression symptomology (44) and Clinician-Administered PTSD Scale (CAPS) for PTSD symptomology (45) used in current proof-of-concept trials. Nonetheless, these measures are important to validate the relationship between psychedelic experience and antidepressant effects, to quantify the degree of MDD and PTSD symptomology that can contribute to substance use, and to track changes in participant quality of life.

Neuroimaging outcomes from clinical trials using psychedelics are highly sought after, since biological data in humans has yet to clearly demonstrate how the acute effects of psychedelics produce clinically relevant and enduring changes in mood and behavior, how dose-dependent 5-HT_2A_ activation contributes to long-term effects, and how long the enduring neuroplastic effects last (46). Further, understanding how maladaptive brain region connectivity and functioning are altered or reversed during and after psychedelic administration is needed to understand how psychedelics may exert lasting therapeutic effects in the clinic. Clinical trials utilizing fMRI (resting-state, active-task, or cue-reactivity) or PET imaging prior to and following psychedelic administration can help answer these questions, as imaging has been the best approach for understanding psychedelic-induced brain changes thus far (47). The increasing demand for these data is evident and nine of the 32 clinical trials described above utilize either fMRI, PET, or MRI (NCT04141501; NCT06405607; NCT06349083; NCT05416229; NCT05421065; NCT05995769; NCT02037126; NCT05322954; NCT06160284) compared to only three trials in 2022 (28). From fMRI, clarity for lasting therapeutic role of psychedelics can be obtained by comparing changes in brain region activity with no task (resting-state) or during a drug-related task, like watching videos related to a drug of choice to measure craving (48), to fMRI data gathered prior to psilocybin administration. To our knowledge, one clinical trial utilizing PET imaging will be the first to measure synaptic density changes in individuals with SUDs before and after psychedelic-assisted psychotherapy (NCT06160284). Synaptic density changes will be measured in subregions of the prefrontal cortex (PFC) that have been implicated by preclinical and preliminary clinical studies in hopes of confirming these results in a clinical setting. This is one of many examples of how clinical and preclinical research can mirror one another to validate findings and create continuity between animal and human research.

### Endpoints and Expected Results

Two important unanswered questions regarding psychedelic-assisted treatments are how rapidly therapeutic changes occur and how long beneficial effects last. Clinical trials attempt to address these gaps through repeated measures over time, through measurement of a variety of endpoints and long-term follow-up. For example, in one clinical trial testing psilocybin in AUD, fMRIs are carried out before, immediately after, and one month following psychedelic administration to compare rapid and lasting changes associated with treatment (NCT04141501). Meanwhile two other clinical trials comparing psilocybin- and ketamine-assisted psychotherapy conduct fMRI before, immediately after (NCT05421065), and one month (NCT05421065) and 3 months (NCT06405607) after treatment. By conducting similar methodology at different endpoints, a greater understanding of lasting effects and differences between endpoints can be achieved. Of the studies described herein, the longest primary endpoint of twelve months is shared between four psychedelic intervention trials for adverse events (NCT06442423) and smoking cessation (NCT01943994; NCT05452772), and one trial of MDMA intervention for PTSD (NCT05709353). There are seven clinical trials with primary endpoints of six months and six clinical trials with primary endpoints of three months with both consisting mostly of TLFB data for substance use, cessation, and abstinence. The shortest primary endpoint is six hours, but the primary outcomes of this clinical trial include safety, and self-reported drug effects (NCT06102434). Therefore, the short primary endpoint is appropriate for the study’s goals and experimental design.

With primary endpoints reaching as far as one year after psychedelic administration and repeated measures often occurring every few weeks or months, clinical trials are adequately addressing questions regarding rapid vs. sustained therapeutic effects of psychedelic treatment. Soon enough, we will have data addressing these concerns to give a better understanding of what changes occur in individuals with SUDs following psychedelic treatment and what the therapeutic potential of this approach might be. Although no results have been posted as of publication of this article, four clinical trials have un-updated, expected, or completed results in 2023 and results are expected for four trials in 2024, 13 trials in 2025, four trials in 2026, four trials in 2027, and one trial in 2029. If data from the first few years of clinical trials remain promising and current trends in research continue, a new wave of psychedelic research could emerge due to the generalizability of psychedelics to other psychiatric disorders that lack efficacious therapies, like SUDs.

### Considerations for future trials: The Mystical Experience, Functional Unblinding, and Application?

Regardless of promising results in clinical trials, there are foundational concerns raised in the FDA’s draft guidance that should be addressed, like limiting the necessity of the mystical experience and subsequent participant functional unblinding (49). Consequently, there is a great desire to establish therapeutic effects in the absence of a mystical experience or using new treatments that lack psychedelic effects (50, 51), which would avert functional unblinding. One approach to address these critiques is through pretreatment with a 5-HT_2A_ antagonist, like ketanserin, which has been shown to not affect neuroplastic consequences of psychedelic administration despite antagonizing 5-HT_2A_ in mice (52). Characterization of 5-HT_2A_ occupancy by ketanserin has recently been established in healthy humans, assisting future trials by preemptively blocking psychedelic effects or preventing adverse effects caused by poor psychedelic experiences (53). A recent case where an individual with treatment-resistant depression accidentally blocked 5-HT_2A_ activation by psilocybin in a clinical trial with trazadone taken the night prior demonstrated that trazadone prevents the hallucinogenic effects of psilocybin but does not interfere with the antidepressant effects (54). Therefore, utilizing pretreatment with a 5-HT_2A_ antagonist could block hallucinogenic effects in clinical trials, which would prevent unblinding, placebo, and subjective effects, while maintaining therapeutic benefits.

Another consideration that has yet to be fully addressed in the current literature is how, if approved, psychedelics will be prescribed to treat SUDs. Although there is extensive literature on the efficacy of psilocybin assisted psychotherapy treatment regiments, some SUDs have already-available treatment options with established efficacy, such as OUD, AUD, or nicotine use disorder (4). Because of this, it is unlikely psychedelics will be applied in the same manner across all SUDs. For SUDs that lack available treatment options, like cocaine use disorder, psilocybin assisted psychotherapy will likely be adopted as a first-line treatment option. On the other hand, for SUDs with currently available treatment options, psychedelics may be more appropriate as an adjunctive treatment option for individuals that are interested, suffer from comorbid conditions, or those with treatment noncompliance or cessation. Ongoing clinical trials are investigating the feasibility of psilocybin as an adjunctive therapy with buprenorphine for OUD (NCT04161066; NCT06067737) and for prescription opioid tapering (NCT05585229). Because current treatment is often discontinued prior to psilocybin administration in clinical trials to directly measure the effectiveness of psilocybin, not much is known about the long-term safety and adjunctive use of psilocybin currently available treatments. Indeed, a recently published case study outlined this very problem for a male individual with comorbid MDD, generalized anxiety disorder, and AUD that relapsed to alcohol use following a cessation of combined escitalopram and naltrexone treatment for a psilocybin experience (55). Therefore, clinicians and researchers should express great caution when utilizing a psychedelic intervention and more research should be done on the safety, efficacy, and feasibility of psychedelics without tapering off current stable treatments.

### Treatment of SUD symptoms using psychedelics: In search of a neurobiological rationale

To appreciate the mechanisms underlying psychedelic effectiveness in treating SUDs, we should first understand the neurobiological underpinnings of SUD symptoms. SUDs are a category of chronic, relapsing conditions and, like many other psychiatric disorders, are characterized by high behavioral and biological complexity. Although our understanding has improved in recent decades, fundamental knowledge about the biology of SUDs is still lacking, which interferes with development of improved approaches for symptom management. Both animal models and human studies have greatly contributed to describing behavioral and neurobiological aspects of SUD, which progresses from an initial rewarding experience of drug consumption (56, 57) in certain settings (58, 59) to chronic, compulsive drug seeking and use despite adverse consequences (60, 61). Within this theoretical framework, animal studies have identified some of the neurobiological mechanisms underlying this progression, many of which are supported by evidence from human patients. However, the descriptions that follow should not be considered paradigmatic of every type of SUD nor of all patients with SUDs.

The early experience of reward following the first exposure to an addictive substance appears to be foundational to the perseverative nature of SUDs and can be considered the starting point of the disease, ultimately leading to pathological, compulsive drug seeking behavior (**Fig 1**). This early stage is characterized neurobiologically by rapid spikes of dopamine release elicited by the addictive substance (56, 62, 63) in the nucleus accumbens (NAc), one of the projection areas from midbrain neurons encoding responses to natural and drug-related rewards and other salient stimuli (64–70). Given that spikes of dopamine in certain portions of the NAc encode rewarding sensations, motor activity, motivation and conditioned associations (71–74), the experience of reward induced by consumption of addictive substances per se is sufficient to trigger continued drug seeking, and to generate an association that elicits subsequent drug seeking and use (71, 75, 76). After repeated drug use, it has been suggested that drug seeking behavior is no longer driven by the experience of reward following consumption but is instead encoded as a habit (77–82) guided in part by environmental and interoceptive cues (83, 84). Accordingly, a prominent theory describing the causes of the progressive escalation of drug seeking suggests that stimuli that are, by nature, present in the environment when drug consumption takes place become increasingly more salient (incentive-sensitization) from a cognitive, motor, and emotional standpoint and become motivating even in the absence of the reward, described as a shift from ‘liking’ to ‘wanting’ the addictive substance (68, 76).

**Figure 1:**
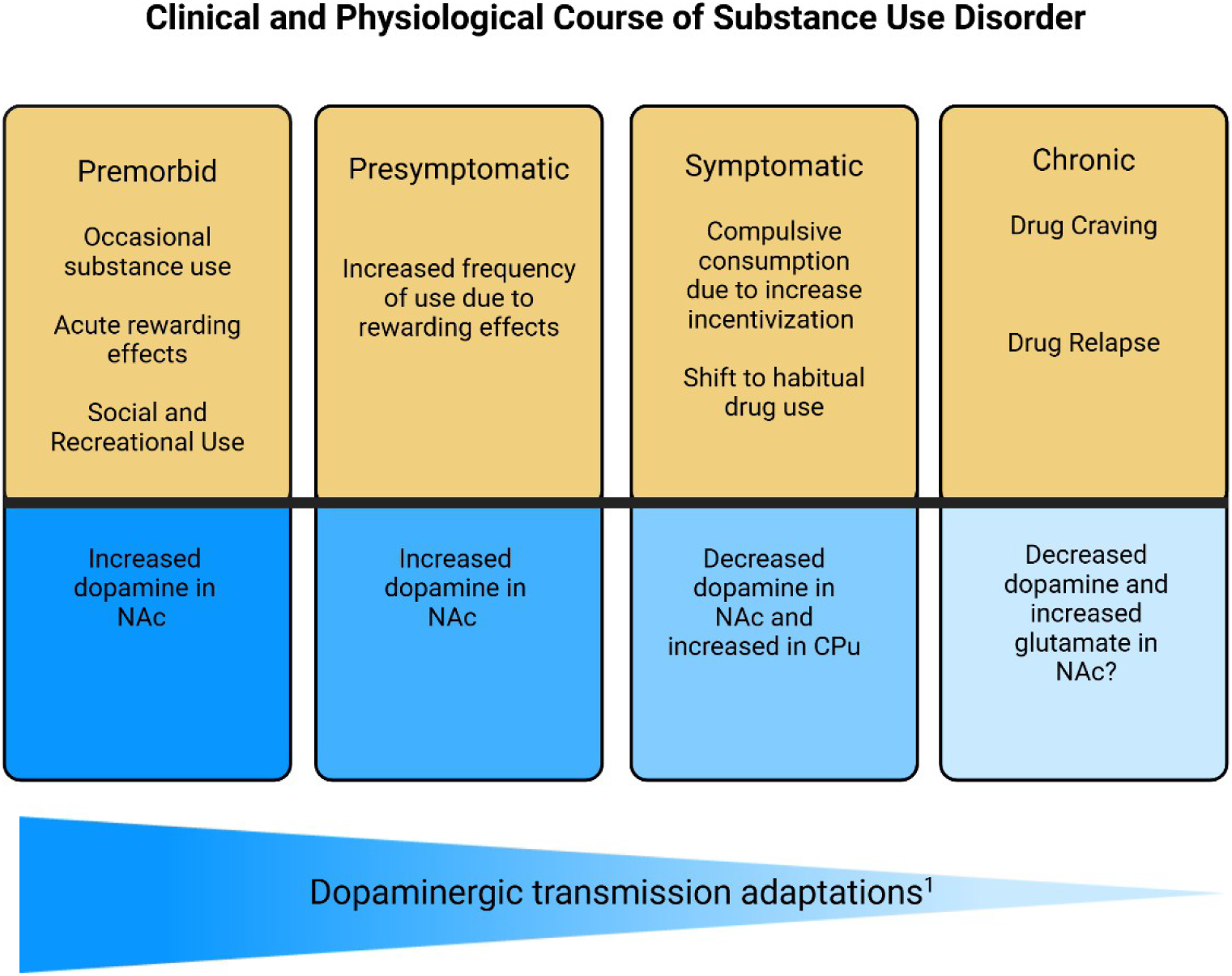
Clinical and Physiological Course of Substance Use Disorder. During premorbid substance use, acute consumption of addictive drugs increases phasic dopamine transmission within the NAc. This transient rewarding experience will motivate a subset of individuals to continue drug use, resulting in progression to a presymptomatic stage where frequency of drug use increases further, increasing phasic episodes of dopamine within the NAc. During the symptomatic stage, individuals shift from reward seeking to compulsive, habitual use, characterized by phasic dopamine decreases in the NAc and increases in the dorsal striatum, particularly the caudate putamen (CPu). As substance use continues, individuals shift to a chronic state where both dopamine and glutamate levels are reduced tonically, permitting phasic increases that drive feelings of craving and lead to drug relapse. Contributions of dysregulated tonic dopamine are relatively unknown at this stage. Please note that withdrawal is not characterized in this figure. ^1^Refers to the extent to which SUD symptoms are dependent upon tonic dopaminergic signaling.

The shift from reward- to cue-driven drug-seeking behavior, whether it results from sensitization, incentive-sensitization or habit, is thought to be influenced by a gradual dominance of the dorsal over the ventral striatum and a gradual shift from reliance on dopamine to a reliance on glutamate signaling in the ventral striatum (85–92). Accordingly, animals with multiple weeks of drug use display high dopamine levels in the dorsal, but not in the ventral striatum (93, 94). Likewise increased dopamine signaling in the dorsal, but not the ventral striatum appears to encode responses to cues associated with addictive drug intake in rodents (93, 95), primates (96, 97), and humans (84, 98); although this dominance of the dorsal over the ventral striatum in response to cues is not always supported by studies in animal models and may depend in part on the behavioral protocols used (99–105). Regardless, during this stage where cues play an important role in driving drug use in SUDs (71, 106, 107), homeostatic changes in dopaminergic transmission appear to be associated with hypofrontality (63) and potentiation of basolateral amygdala activity (108–110).

During withdrawal from chronic drug use, transcriptional, epigenetic (111), and physiological (88, 112–116) changes occur that result in persistent structural and functional alterations in neurons and astrocytes within the cortico-limbic-striatal network. Indeed, the literature strongly supports a role for altered excitatory transmission in regulating craving and relapse to drug use during this stage (77, 82, 85–87, 90, 91, 110, 117), alongside reduced dopamine transmission within the entire striatum (63). Taken together, it can be understood that dysfunctional dopamine homeostasis within the ventral striatum initiates SUD development, while dysfunctional glutamate homeostasis in response to changes in dopamine neurotransmission are critical for the maintenance of SUDs and relapse.

Despite our vast knowledge of SUDs, it is not yet clear how psychedelics work within this framework to produce therapeutic benefits. In humans, psychedelics alter functional connectivity of brain regions within the default mode network (DMN) (118), which are commonly altered in SUDs (119). The DMN is a group of brain regions that support emotional processing, mental activity, and memory recollection through involvement of brain regions including the PFC, cingulate cortex, hippocampus, and parietal and temporal cortices (119). DMN dysfunction is observed across multiple drug classes and is associated with drug addiction (120), craving (121), and relapse (122), as well as comorbid diagnoses such as MDD (123) and PTSD (124). Following psilocybin administration, deficits in thalamus and cingulate cortex functioning (125) and functional connectivity between medial PFC- posterior cingulate cortex and DMN-anterior hippocampus (126) are observed. Psilocybin also appears to have a “correction effect” as administration to individuals diagnosed with treatment-resistant depression, who typically lack responsiveness, have increased amygdala responsiveness to emotional stimuli (127, 128), while healthy volunteers experience decreased amygdala responsiveness (129). The molecular mechanisms underlying these changes remain unclear. Currently, it is believed that psychedelics stimulate neuroplasticity through activation of PFC pyramidal neurons that project to the ventral tegmental area (VTA), NAc, and amygdala (130). When administered alongside standardized behavioral treatment regimens like psychotherapy, psychedelic-induced neuroplasticity has the potential to rewire maladaptive brain circuitry caused by SUDs.

The primary mechanism of action for classical psychedelics is through 5-HT_2A_ agonism (**Fig. 2**). Activation of 5-HT_2A_ receptors causes hallmark symptoms, like hallucinations and delusions (131–133) and activates PLC-β-triggered cascades for protein kinase C (PKC) and release of intracellular Ca^2+^ stores for calcium/calmodulin kinase II (CaMKII) to promote synaptic plasticity (134, 135). PKC activation induces trafficking of excitatory AMPA and NMDA receptors to the postsynaptic membrane (136), and CaMKII induces Ca^2+^ signaling at glutamatergic synapses supporting long-term potentiation (LTP) (137). Recently, it has been reported that activation of internalized 5-HT_2A_ receptors by DMT, a membrane-permeable ligand, induces dendritic spine generation and antidepressant effects and can explain why other ligands, such as serotonin, cannot induce similar neuroplasticity (138).

**Figure 2:**
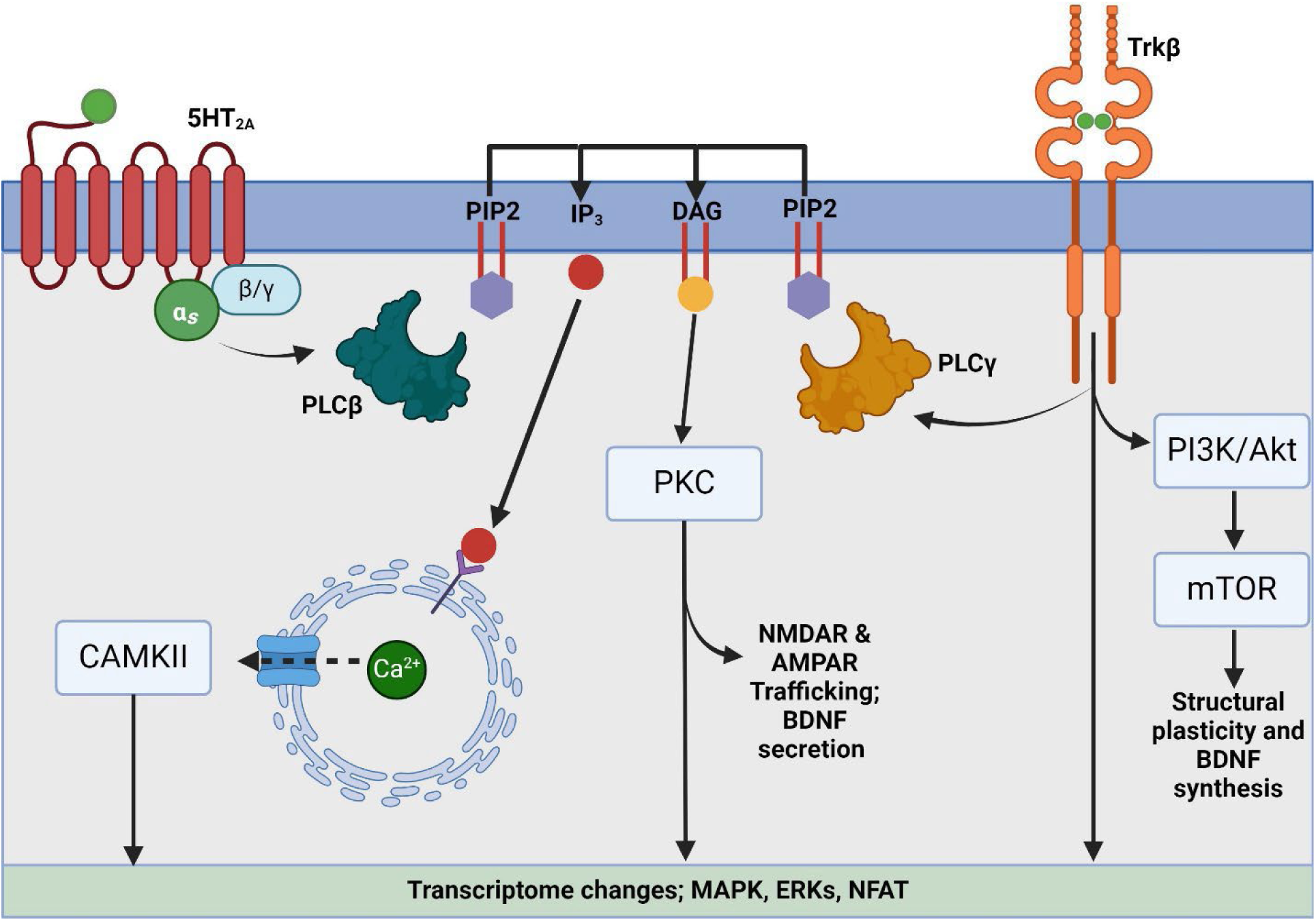
Signaling cascades underlying psychedelic-induced plasticity. Classical psychedelics bind directly to 5HT_2A_ receptors and allosterically to Trkβ receptors causing a conformational change in PLCβ and PLCγ, respectively, allowing for activation of PIP2. PIP2 then activates IP_3_ causing downstream intracellular release of calcium, production of CAMKII, and transcriptome changes. Hydrolysis of PIP2 leads to production of DAG, which activates PKC resulting in NMDAR and AMPAR trafficking to the cell membrane, BDNF secretion, and transcriptome changes. Trkβ activation directly causes transcriptome changes via MAPK and activates the PI3K-Akt-mTOR pathway for synthesis of proteins involved in structural plasticity, including BDNF.

Apart from 5HT_2A_ agonism, psychedelics can induce neuroplasticity through allosteric binding to Trkβ receptors that prolong dimerization for brain derived neurotrophic factor (BDNF) binding (139).

Once bound, Trkβ receptors activate multiple signaling cascades including PI3K-Akt for neuronal development, synapse formation, and neuroprotection (140, 141), MAPK for learning, memory, early response genes associated with plasticity (142), and PLC-γ for Ca^2+^ release (143) in a similar manner as 5HT_2A_ activation. Interestingly, BDNF activation of Trkβ is hypothesized to induce a positive feedback loop by activating target of rapamycin (mTOR), which in turn increases BDNF synthesis (144) that, alongside sustained AMPA receptor activation for BDNF secretion, could then further activate Trkβ (145). This cycle could explain the neuroplastic effects that are observed in clinical populations long after the elimination of the psychedelic.

Another potential therapeutic mechanism of action of psychedelics is through dopamine restoration within the NAc. As discussed previously, the initial stages of substance use is dictated by phasic dopamine release induced by addictive substances (56, 62, 63) in the NAc that can lead to the development of a SUD and dysregulation of dopamine homeostasis (**Fig. 1**). Indeed, striatal dopamine release is blunted by chronic drug use (146–148), which has been suggested to predict poor treatment response and increased vulnerability to SUDs (149). Therefore, striatal dopamine restoration alongside behavioral treatment could be therapeutic for SUDs (150). Although data is limited, it is known that pscyhedelics, particularly psilocybin and LSD, are capable of increasing striatal dopamine (151). Physiological levels of dopamine within the striatum are necessary to achieve therapeutic outcomes in psychiatric conditions (152–155). Accordingly, mild stimulation of dopamine release is effective in reducing alcohol seeking in animal models (156) and current FDA-approved treatment for nicotine use disorder using nicotine replacement therapy, bupropion, or other antidepressants (157), work as surrogates of dopamine to stimulate the reward system and reduce the need for nicotine consumption (158–160). Therefore, we became interested in the consequential dopaminergic output within the NAc following psychedelic administration to better understand how psychedelics are mechanistically and neurobiologically therapeutic for SUDs.

To validate the potential of psychedelics to function via dopamine restoration, we systematically identified literature that utilized *in vivo* microdialysis in rats to measure dopamine concentrations within the NAc following administration of a psychedelic or MDMA. Literature searches were conducted using PubMed and SCOPUS in August 2024 without date or language restrictions for studies published using *in* vivo microdialysis to detect extracellular dopamine changes within the nucleus accumbens following administration of a psychedelic. Grey literature searches were conducted using Google Scholar. Most identified studies utilized wild-type rat, so manuscripts using mice were excluded for potential species differences. Data was extracted and quantified with WebPlotDigitizer and GraphPad Prizm 10 software (**Fig. 3**). Following drug administration, there was an immediate increase in dopamine for all experimental treatments, especially for high-dose MDMA. Interestingly, animals that received 10mg/kg psilocybin had a sustained increase in extracellular dopamine (+ ∼50%) throughout the experiment while both MDMA doses returned dopamine concentrations to vehicle levels. Notably, high-dose MDMA (5mg/kg) -induced dopamine increases peaks much higher than psilocybin (+ ∼250%), suggesting that interventions of high dose MDMA could be less effective or deleterious for individuals with SUD. Though the increase in dopamine release stimulated by psilocybin would raise doubts about their potential abuse liability, in reality, psilocybin stimulates approximately 1/10^th^ of the dopamine released by psychostimulants like cocaine (165), amphetamine (166), and methamphetamine (167), thus supporting the low prevalence of psychedelic abuse (132, 168). Further, psychostimulant-induced dopamine stimulation rapidly peaks at levels as high as a 1200% of baseline and decays to levels as high as 300% over the course of seven hours (166), while psilocybin sustains a much milder and long-lasting increase (∼50%) in dopamine levels (**Fig. 3**). This different impact on dopamine release elicited by psychostimulants and psychedelics appears to indicate that psychostimulants preferentially potentiate phasic dopamine release, which is thought to underlie abuse liability of addictive drugs (104, 169–171), whereas psilocybin prominently activate tonic release of dopamine, which has inhibitory effects on both neurotransmission within striatal circuitries and behavior (152–155) Obviously, it is difficult at this point to attribute the effects of psychedelics entirely to tonic and phasic dopamine transmission, since the net amount of extracellular dopamine detected during microdialysis results from release and uptake dynamics affecting the rate transport of analytes between extracellular and intracellular compartments (172). Thus, while clarity on these points requires future studies, the sustained elevation of tonic dopamine release is consistent with the dopamine-restoration theory of psychedelics in SUDs, although more work needs to be done to fully understand this mechanism. Assessments of extracellular dopamine levels at extended time points after psychedelic administration and during addictive drug self-administration and withdrawal could determine the validity of the dopamine-restoration theory as a potential therapeutic mechanism by which psychedelics are efficacious in treatment of SUDs.

**Figure 3:**
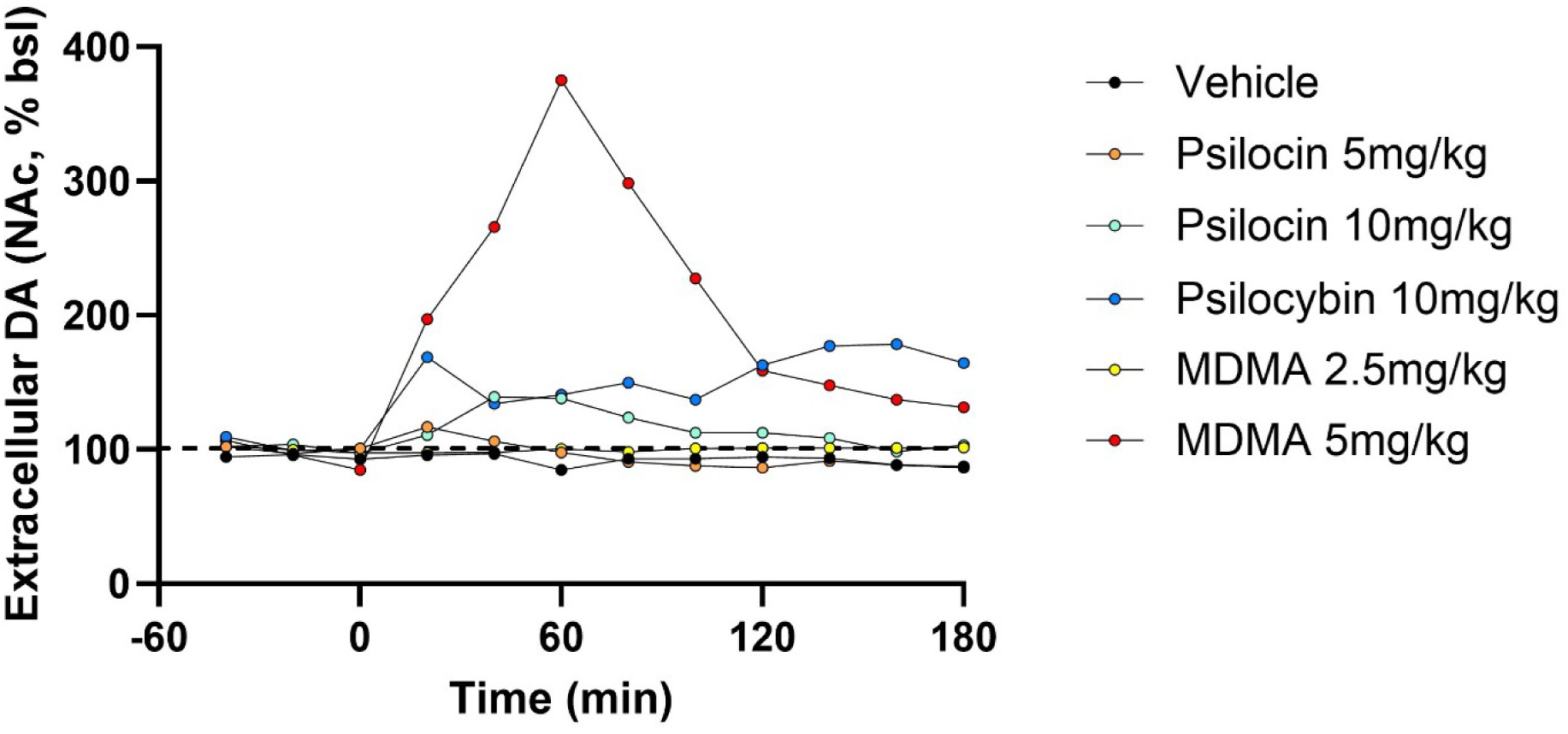
Psilocybin and MDMA increase extracellular dopamine within the NAc. Psilocybin (10mg/kg), but not psilocin, induces a rapid increase of accumbal dopamine that is prolonged (∼50%) for three hours. Psilocin (10mg/kg) causes a temporary increase of accumbal dopamine, but is not sustained. MDMA (5mg/kg), but not MDMA (2.5mg/kg), increases accumbal dopamine with a peak 60 minutes after administration. <u>Psilocybin (161, 162) and MDMA (163, 164) data were extracted, adapted, and modified to 180 minutes for uniformity.</u>

### Limitations

In this paper, we conducted two independent systematic literature searches to characterize currently registered clinical trials and to summarize *in vivo* microdialysis data within the nucleus accumbens of rats from published manuscripts. In our characterization of clinical trials, it is possible that clinical trials were missed due to being outside of the scope of our search terms. For example, clinical trials could report a common comorbid condition of SUDs, like the three trials in Table 1 also studying PTSD (NCT05219175, NCT05943665, and NCT05709353), that may not list SUDs as the condition of interest despite measuring SUD symptomology. Further, because all searches were conducted on ClinicalTrials.gov, it is possible that some clinical trials under other registries, such as the International Clinical Trials Registry Platform (ICTRP), could have been missed and remained uncharacterized.

To summarize *in vivo* microdialysis data, we conducted searches only on PubMed, SCOPUS, and Google Scholar search engines, which could cause some publications to remain undetected. Further, due to the novelty of psychedelics affecting dopamine in reward circuitry, only three publications met our criteria (Supp. Fig. 2; Supp. Table 1) and only measured psilocybin or MDMA (**Fig. 3**). Therefore, more studies utilizing a range of doses and the inclusion of other classic psychedelics are needed to determine if mild dopamine stimulation is psilocybin- or psychedelic-specific. Another limitation to this work is that SUD is complex involving many brain regions, pathways, and neurotransmitters. While our focus was only on dopamine stimulation within the nucleus accumbens, a foundational element to development and symptomology of SUDs, it is important to acknowledge this mechanism likely exists with other alterations in brain function and signaling to manifest symptoms of SUDs and the therapeutic potential of psychedelics for diagnoses that involve reward circuitry malfunction.

## Conclusion

With resurging interest in psychedelics as a pharmacotherapy, the field is rapidly changing as researchers work to find better treatments for various mood and behavioral disorders. While work remains to be done to fully understand the therapeutic potential of psychedelics, great strides have been made thus far in both animal and human research to probe the underlying neurobiological mechanisms of these processes and extent to which they last. Herein, we described ongoing clinical trials registered with ClinicalTrials.gov investigating psychedelics for SUDs, provide insight into how psychedelics may be therapeutic within a SUD framework, and propose dopamine restoration via tonic dopamine stimulation in the nucleus accumbens as a novel neurobiological mechanism that may contribute to the success of psychedelics in clinical trials. Future preclinical and clinical work is needed to further validate and characterize this mechanism for SUDs with the potential to explain therapeutic effects of psychedelics mechanistically and change the current framework for how we currently treat SUDs.

## Data Availability

All data produced in the present study are available upon reasonable request to the authors.

## Acknowledgments

The authors would like to thank Dr. Anna Kruyer for reading the manuscript and providing constructive feedback.

## References

1. Administration SAaMHS. Key substance use and mental health indicators in the United States: Results from the 2021 National Survey on Drug Use and Health. Center for Behavioral Health Statistics and Quality; 2022.

2. Li M, Peterson C, Xu L, Mikosz CA, Luo F. Medical Costs of Substance Use Disorders in the US Employer-Sponsored Insurance Population. JAMA Netw Open. 2023;6(1):e2252378.

3. NIDA. Treatment and Recovery 2023 [Available from: https://nida.nih.gov/publications/drugs-brains-behavior-science-addiction/treatment-recovery.

4. Volkow ND. Personalizing the Treatment of Substance Use Disorders. Am J Psychiatry. 2020;177(2):113–6.

5. Muttoni S, Ardissino M, John C. Classical psychedelics for the treatment of depression and anxiety: A systematic review. J Affect Disord. 2019;258:11–24.

6. Nichols DE, Johnson MW, Nichols CD. Psychedelics as Medicines: An Emerging New Paradigm. Clin Pharmacol Ther. 2017;101(2):209–19.

7. Nutt D, Carhart-Harris R. The Current Status of Psychedelics in Psychiatry. JAMA Psychiatry. 2021;78(2):121–2.

8. Reiff CM, Richman EE, Nemeroff CB, Carpenter LL, Widge AS, Rodriguez CI, et al. Psychedelics and Psychedelic-Assisted Psychotherapy. Am J Psychiatry. 2020;177(5):391–410.

9. Tricco AC, Lillie E, Zarin W, O’Brien KK, Colquhoun H, Levac D, et al. PRISMA Extension for Scoping Reviews (PRISMA-ScR): Checklist and Explanation. Ann Intern Med. 2018;169(7):467–73.

10. Savage C, McCabe OL. Residential psychedelic (LSD) therapy for the narcotic addict. A controlled study. Arch Gen Psychiatry. 1973;28(6):808–14.

11. Fuentes JJ, Fonseca F, Elices M, Farre M, Torrens M. Therapeutic Use of LSD in Psychiatry: A Systematic Review of Randomized-Controlled Clinical Trials. Front Psychiatry. 2019;10:943.

12. Grinspoon L, Bakalar JB. Psychedelic drugs reconsidered. New York: Basic Books; 1979.

13. Gable RS. Comparison of acute lethal toxicity of commonly abused psychoactive substances. Addiction. 2004;99(6):686–96.

14. Nichols DE, Grob CS. Is LSD toxic? Forensic Sci Int. 2018;284:141–5.

15. de Veen BT, Schellekens AF, Verheij MM, Homberg JR. Psilocybin for treating substance use disorders? Expert Rev Neurother. 2017;17(2):203–12.

16. Worrell SD, Gould TJ. Therapeutic potential of ketamine for alcohol use disorder. Neurosci Biobehav Rev. 2021;126:573–89.

17. Fitzgerald ND, Striley CW, Palamar JJ, Copeland J, Kurtz S, Cottler LB. Test-retest reliability and cross-cultural applicability of DSM-5 adopted diagnostic criteria for ketamine use disorders. Drug Alcohol Depend. 2021;228:109056.

18. Hofmann A. Psychotomimetic drugs: Chemical and Pharmacological Aspects. Acta Physiologica et Pharmacologica Neerlandica. 1959;8:240–58.

19. FDA grants Breakthrough Therapy Designation to Usona Institute’s psilocybin program for major depressive disorder [press release]. 2019.

20. Davis AK, Barrett FS, May DG, Cosimano MP, Sepeda ND, Johnson MW, et al. Effects of Psilocybin-Assisted Therapy on Major Depressive Disorder: A Randomized Clinical Trial. JAMA Psychiatry. 2021;78(5):481–9.

21. Raison CL, Sanacora G, Woolley J, Heinzerling K, Dunlop BW, Brown RT, et al. Single-Dose Psilocybin Treatment for Major Depressive Disorder: A Randomized Clinical Trial. JAMA. 2023;330(9):843–53.

22. von Rotz R, Schindowski EM, Jungwirth J, Schuldt A, Rieser NM, Zahoranszky K, et al. Single-dose psilocybin-assisted therapy in major depressive disorder: A placebo-controlled, double-blind, randomised clinical trial. EClinicalMedicine. 2023;56:101809.

23. Gukasyan N, Davis AK, Barrett FS, Cosimano MP, Sepeda ND, Johnson MW, et al. Efficacy and safety of psilocybin-assisted treatment for major depressive disorder: Prospective 12-month follow-up. J Psychopharmacol. 2022;36(2):151–8.

24. Bogenschutz MP, Forcehimes AA, Pommy JA, Wilcox CE, Barbosa PC, Strassman RJ. Psilocybin-assisted treatment for alcohol dependence: a proof-of-concept study. J Psychopharmacol. 2015;29(3):289–99.

25. Johnson MW, Garcia-Romeu A, Cosimano MP, Griffiths RR. Pilot study of the 5-HT2AR agonist psilocybin in the treatment of tobacco addiction. J Psychopharmacol. 2014;28(11):983–92.

26. Johnson MW, Garcia-Romeu A, Griffiths RR. Long-term follow-up of psilocybin-facilitated smoking cessation. Am J Drug Alcohol Abuse. 2017;43(1):55–60.

27. Bogenschutz MP, Ross S, Bhatt S, Baron T, Forcehimes AA, Laska E, et al. Percentage of Heavy Drinking Days Following Psilocybin-Assisted Psychotherapy vs Placebo in the Treatment of Adult Patients With Alcohol Use Disorder: A Randomized Clinical Trial. JAMA Psychiatry. 2022;79(10):953–62.

28. van der Meer PB, Fuentes JJ, Kaptein AA, Schoones JW, de Waal MM, Goudriaan AE, et al. Therapeutic effect of psilocybin in addiction: A systematic review. Front Psychiatry. 2023;14:1134454.

29. de la Torre R, Farre M, Roset PN, Pizarro N, Abanades S, Segura M, et al. Human pharmacology of MDMA: pharmacokinetics, metabolism, and disposition. Ther Drug Monit. 2004;26(2):137–44.

30. Brown TK. Ibogaine in the treatment of substance dependence. Curr Drug Abuse Rev. 2013;6(1):3–16.

31. Sessa B. MDMA and PTSD treatment: "PTSD: From novel pathophysiology to innovative therapeutics". Neurosci Lett. 2017;649:176–80.

32. Holma M, Holma I, Isometsa E. Comorbid alcohol use disorder in psychiatric MDD patients: A five-year prospective study. J Affect Disord. 2020;267:283–8.

33. Hunt GE, Malhi GS, Lai HMX, Cleary M. Prevalence of comorbid substance use in major depressive disorder in community and clinical settings, 1990-2019: Systematic review and meta-analysis. J Affect Disord. 2020;266:288-304.

34. Johnson MW. Classic Psychedelics in Addiction Treatment: The Case for Psilocybin in Tobacco Smoking Cessation. Curr Top Behav Neurosci. 2022;56:213–27.

35. Stolberg HO, Norman G, Trop I. Randomized controlled trials. AJR Am J Roentgenol. 2004;183(6):1539–44.

36. Deaton A, Cartwright N. Understanding and misunderstanding randomized controlled trials. Soc Sci Med. 2018;210:2–21.

37. D’Souza DC, Syed SA, Flynn LT, Safi-Aghdam H, Cozzi NV, Ranganathan M. Exploratory study of the dose-related safety, tolerability, and efficacy of dimethyltryptamine (DMT) in healthy volunteers and major depressive disorder. Neuropsychopharmacology. 2022;47(10):1854–62.

38. Goodwin GM, Aaronson ST, Alvarez O, Arden PC, Baker A, Bennett JC, et al. Single-Dose Psilocybin for a Treatment-Resistant Episode of Major Depression. N Engl J Med. 2022;387(18):1637–48.

39. Barrett FS, Johnson MW, Griffiths RR. Validation of the revised Mystical Experience Questionnaire in experimental sessions with psilocybin. J Psychopharmacol. 2015;29(11):1182–90.

40. Griffiths RR, Johnson MW, Richards WA, Richards BD, McCann U, Jesse R. Psilocybin occasioned mystical-type experiences: immediate and persisting dose-related effects. Psychopharmacology (Berl). 2011;218(4):649–65.

41. Sobell LC, Sobell MB. Timeline Follow-Back. In: Z. LR, Allen JP, editors. Measuring Alcohol Consumption. Totowa, NJ: Humana Press; 1992. p. 41-72.

42. Hjorthoj CR, Hjorthoj AR, Nordentoft M. Validity of Timeline Follow-Back for self-reported use of cannabis and other illicit substances--systematic review and meta-analysis. Addict Behav. 2012;37(3):225–33.

43. Lane SD, Yoon JH, Heads AM, de Dios CI, Yammine L, Hong J, et al. Biomarkers in Substance Use Disorder. In: Teixeira A, Rocha N, Berk M, editors. Biomarkers in Neuropsychiatry. 1 ed: Springer Cham; 2023. p. 291-328.

44. Haikazian S, Chen-Li DCJ, Johnson DE, Fancy F, Levinta A, Husain MI, et al. Psilocybin-assisted therapy for depression: A systematic review and meta-analysis. Psychiatry Res. 2023;329:115531.

45. Smith KW, Sicignano DJ, Hernandez AV, White CM. MDMA-Assisted Psychotherapy for Treatment of Posttraumatic Stress Disorder: A Systematic Review With Meta-Analysis. J Clin Pharmacol. 2022;62(4):463–71.

46. Wall MB, Harding R, Zafar R, Rabiner EA, Nutt DJ, Erritzoe D. Neuroimaging in psychedelic drug development: past, present, and future. Mol Psychiatry. 2023;28(9):3573–80.

47. Nichols DE. Psilocybin: from ancient magic to modern medicine. J Antibiot (Tokyo). 2020;73(10):679–86.

48. Kober H, Lacadie CM, Wexler BE, Malison RT, Sinha R, Potenza MN. Brain Activity During Cocaine Craving and Gambling Urges: An fMRI Study. Neuropsychopharmacology. 2016;41(2):628–37.

49. Research FaDA-CfDEa. Psychedelic Drugs: Considerations for Clinical Investigations Guidance for Industry (Draft Guidance). In: Services USDoHaH, editor.: Office of Communications, Dvision of Drug Information; 2023.

50. Hashimoto K. Are "mystical experiences" essential for antidepressant actions of ketamine and the classic psychedelics? Eur Arch Psychiatry Clin Neurosci. 2024.

51. Olson DE. The Subjective Effects of Psychedelics May Not Be Necessary for Their Enduring Therapeutic Effects. ACS Pharmacol Transl Sci. 2021;4(2):563–7.

52. Hesselgrave N, Troppoli TA, Wulff AB, Cole AB, Thompson SM. Harnessing psilocybin: antidepressant-like behavioral and synaptic actions of psilocybin are independent of 5-HT2R activation in mice. Proc Natl Acad Sci U S A. 2021;118(17).

53. Holze F, Madsen MK, Svarer C, Gillings N, Stenbaek DS, Rudin D, et al. Ketanserin exhibits dose- and concentration-proportional serotonin 2A receptor occupancy in healthy individuals: Relevance for psychedelic research. Eur Neuropsychopharmacol. 2024;88:43–8.

54. Rosenblat JD, Leon-Carlyle M, Ali S, Husain MI, McIntyre RS. Antidepressant Effects of Psilocybin in the Absence of Psychedelic Effects. Am J Psychiatry. 2023;180(5):395–6.

55. Frye MA, Singh B, Breitinger SA, Oesterle TS. Selective Serotonin Reuptake Inhibitor Discontinuation for Psilocybin Treatment and Contributions to Alcohol Addiction Relapse: A Cautionary Tale. J Clin Psychiatry. 2024;85(3).

56. Di Chiara G, Imperato A. Drugs abused by humans preferentially increase synaptic dopamine concentrations in the mesolimbic system of freely moving rats. Proc Natl Acad Sci U S A. 1988;85(14):5274–8.

57. Wise RA, Bozarth MA. A psychomotor stimulant theory of addiction. Psychol Rev. 1987;94(4):469–92.

58. Muller CP, Schumann G. Drugs as instruments: a new framework for non-addictive psychoactive drug use. Behav Brain Sci. 2011;34(6):293–310.

59. Stewart J, Badiani A. Tolerance and sensitization to the behavioral effects of drugs. Behav Pharmacol. 1993;4(4):289–312.

60. American Psychiatric A. Diagnostic and statistical manual of mental disorders: DSM-5. [DSM V]. 5th edition ed. Washington, DC: American Psychiatric Publishing; 2013.

61. Everitt BJ, Robbins TW. Drug Addiction: Updating Actions to Habits to Compulsions Ten Years On. Annu Rev Psychol. 2016;67:23–50.

62. Phillips PE, Stuber GD, Heien ML, Wightman RM, Carelli RM. Subsecond dopamine release promotes cocaine seeking. Nature. 2003;422(6932):614-8.

63. Volkow ND, Fowler JS, Wang GJ, Swanson JM, Telang F. Dopamine in drug abuse and addiction: results of imaging studies and treatment implications. Arch Neurol. 2007;64(11):1575–9.

64. Brischoux F, Chakraborty S, Brierley DI, Ungless MA. Phasic excitation of dopamine neurons in ventral VTA by noxious stimuli. Proc Natl Acad Sci U S A. 2009;106(12):4894–9.

65. Gray JA, Kumari V, Lawrence N, Young AMJ. Functions of the dopaminergic innervation of the nucleus accumbens. Psychobiology. 1999;27:225–35.

66. Horvitz JC. Mesolimbocortical and nigrostriatal dopamine responses to salient non-reward events. Neuroscience. 2000;96(4):651–6.

67. Ikemoto S, Panksepp J. The role of nucleus accumbens dopamine in motivated behavior: a unifying interpretation with special reference to reward-seeking. Brain Res Brain Res Rev. 1999;31(1):6–41.

68. Robinson TE, Berridge KC. The neural basis of drug craving: an incentive-sensitization theory of addiction. Brain Res Brain Res Rev. 1993;18(3):247–91.

69. Salamone JD, Cousins MS, McCullough LD, Carriero DL, Berkowitz RJ. Nucleus accumbens dopamine release increases during instrumental lever pressing for food but not free food consumption. Pharmacol Biochem Behav. 1994;49(1):25–31.

70. Schultz W, Dayan P, Montague PR. A neural substrate of prediction and reward. Science. 1997;275(5306):1593-9.

71. Di Chiara G. Drug addiction as dopamine-dependent associative learning disorder. Eur J Pharmacol. 1999;375(1-3):13–30.

72. Floresco SB, Magyar O. Mesocortical dopamine modulation of executive functions: beyond working memory. Psychopharmacology (Berl). 2006;188(4):567–85.

73. Mogenson GJ, Jones DL, Yim CY. From motivation to action: functional interface between the limbic system and the motor system. Prog Neurobiol. 1980;14(2-3):69–97.

74. Wise RA. Dopamine, learning and motivation. Nat Rev Neurosci. 2004;5(6):483–94.

75. Kelley AE. Neural integrative activities of nucleus accumbens subregions in relation to learning and motivation. Psychobiology. 1999;27:198–213.

76. Robinson TE, Berridge KC. The psychology and neurobiology of addiction: an incentive-sensitization view. Addiction. 2000;95 Suppl 2:S91–117.

77. Belin D, Everitt BJ. Cocaine seeking habits depend upon dopamine-dependent serial connectivity linking the ventral with the dorsal striatum. Neuron. 2008;57(3):432–41.

78. Deroche-Gamonet V, Belin D, Piazza PV. Evidence for addiction-like behavior in the rat. Science. 2004;305(5686):1014-7.

79. Di Ciano P, Everitt BJ. Conditioned reinforcing properties of stimuli paired with self-administered cocaine, heroin or sucrose: implications for the persistence of addictive behaviour. Neuropharmacology. 2004;47 Suppl 1:202–13.

80. Everitt BJ, Dickinson A, Robbins TW. The neuropsychological basis of addictive behaviour. Brain Res Brain Res Rev. 2001;36(2-3):129–38.

81. Hyman SE, Malenka RC. Addiction and the brain: the neurobiology of compulsion and its persistence. Nat Rev Neurosci. 2001;2(10):695–703.

82. Vanderschuren LJ, Everitt BJ. Drug seeking becomes compulsive after prolonged cocaine self-administration. Science. 2004;305(5686):1017-9.

83. Vanderschuren LJ, Di Ciano P, Everitt BJ. Involvement of the dorsal striatum in cue-controlled cocaine seeking. J Neurosci. 2005;25(38):8665–70.

84. Volkow ND, Wang GJ, Telang F, Fowler JS, Logan J, Childress AR, et al. Cocaine cues and dopamine in dorsal striatum: mechanism of craving in cocaine addiction. J Neurosci. 2006;26(24):6583–8.

85. Berke JD, Hyman SE. Addiction, dopamine, and the molecular mechanisms of memory. Neuron. 2000;25(3):515–32.

86. Everitt BJ, Robbins TW. Neural systems of reinforcement for drug addiction: from actions to habits to compulsion. Nat Neurosci. 2005;8(11):1481–9.

87. Everitt BJ, Wolf ME. Psychomotor stimulant addiction: a neural systems perspective. J Neurosci. 2002;22(9):3312–20.

88. Kalivas PW. The glutamate homeostasis hypothesis of addiction. Nat Rev Neurosci. 2009;10(8):561–72.

89. Robbins TW, Everitt BJ. Drug addiction: bad habits add up. Nature. 1999;398(6728):567-70.

90. Schneck N, Vezina P. Enhanced dorsolateral striatal activity in drug use: the role of outcome in stimulus-response associations. Behav Brain Res. 2012;235(2):136–42.

91. White NM. Addictive drugs as reinforcers: multiple partial actions on memory systems. Addiction. 1996;91(7):921–49; discussion 51-65.

92. Everitt BJ, Robbins TW. From the ventral to the dorsal striatum: devolving views of their roles in drug addiction. Neurosci Biobehav Rev. 2013;37(9 Pt A):1946-54.

93. Ito R, Dalley JW, Howes SR, Robbins TW, Everitt BJ. Dissociation in conditioned dopamine release in the nucleus accumbens core and shell in response to cocaine cues and during cocaine-seeking behavior in rats. J Neurosci. 2000;20(19):7489–95.

94. Ito R, Dalley JW, Robbins TW, Everitt BJ. Dopamine release in the dorsal striatum during cocaine-seeking behavior under the control of a drug-associated cue. J Neurosci. 2002;22(14):6247–53.

95. Neisewander JL, O’Dell LE, Tran-Nguyen LT, Castaneda E, Fuchs RA. Dopamine overflow in the nucleus accumbens during extinction and reinstatement of cocaine self-administration behavior. Neuropsychopharmacology. 1996;15(5):506–14.

96. Letchworth SR, Nader MA, Smith HR, Friedman DP, Porrino LJ. Progression of changes in dopamine transporter binding site density as a result of cocaine self-administration in rhesus monkeys. J Neurosci. 2001;21(8):2799–807.

97. Porrino LJ, Lyons D, Smith HR, Daunais JB, Nader MA. Cocaine self-administration produces a progressive involvement of limbic, association, and sensorimotor striatal domains. J Neurosci. 2004;24(14):3554–62.

98. Garavan H, Pankiewicz J, Bloom A, Cho JK, Sperry L, Ross TJ, et al. Cue-induced cocaine craving: neuroanatomical specificity for drug users and drug stimuli. Am J Psychiatry. 2000;157(11):1789–98.

99. Boileau I, Dagher A, Leyton M, Welfeld K, Booij L, Diksic M, et al. Conditioned dopamine release in humans: a positron emission tomography [11C]raclopride study with amphetamine. J Neurosci. 2007;27(15):3998–4003.

100. Jasinska AJ, Stein EA, Kaiser J, Naumer MJ, Yalachkov Y. Factors modulating neural reactivity to drug cues in addiction: a survey of human neuroimaging studies. Neurosci Biobehav Rev. 2014;38:1–16.

101. Leyton M, Vezina P. On cue: striatal ups and downs in addictions. Biol Psychiatry. 2012;72(10):e21–2.

102. Singer BF, Fadanelli M, Kawa AB, Robinson TE. Are Cocaine-Seeking "Habits" Necessary for the Development of Addiction-Like Behavior in Rats? J Neurosci. 2018;38(1):60–73.

103. Vollstadt-Klein S, Wichert S, Rabinstein J, Buhler M, Klein O, Ende G, et al. Initial, habitual and compulsive alcohol use is characterized by a shift of cue processing from ventral to dorsal striatum. Addiction. 2010;105(10):1741–9.

104. Willuhn I, Burgeno LM, Everitt BJ, Phillips PE. Hierarchical recruitment of phasic dopamine signaling in the striatum during the progression of cocaine use. Proc Natl Acad Sci U S A. 2012;109(50):20703–8.

105. Willuhn I, Burgeno LM, Groblewski PA, Phillips PE. Excessive cocaine use results from decreased phasic dopamine signaling in the striatum. Nat Neurosci. 2014;17(5):704–9.

106. Childress AR, McLellan AT, Ehrman R, O’Brien CP. Classically conditioned responses in opioid and cocaine dependence: a role in relapse? NIDA Res Monogr. 1988;84:25–43.

107. O’Brien CP, Childress AR, McLellan AT, Ehrman R. A learning model of addiction. Res Publ Assoc Res Nerv Ment Dis. 1992;70:157–77.

108. Cador M, Robbins TW, Everitt BJ. Involvement of the amygdala in stimulus-reward associations: interaction with the ventral striatum. Neuroscience. 1989;30(1):77–86.

109. Meil WM, See RE. Lesions of the basolateral amygdala abolish the ability of drug associated cues to reinstate responding during withdrawal from self-administered cocaine. Behav Brain Res. 1997;87(2):139–48.

110. Whitelaw RB, Markou A, Robbins TW, Everitt BJ. Excitotoxic lesions of the basolateral amygdala impair the acquisition of cocaine-seeking behaviour under a second-order schedule of reinforcement. Psychopharmacology (Berl). 1996;127(3):213–24.

111. Robison AJ, Nestler EJ. Transcriptional and epigenetic mechanisms of addiction. Nat Rev Neurosci. 2011;12(11):623–37.

112. Bonci A, Williams JT. Increased probability of GABA release during withdrawal from morphine. J Neurosci. 1997;17(2):796–803.

113. Kruyer A, Dixon D, Angelis A, Amato D, Kalivas PW. Astrocytes in the ventral pallidum extinguish heroin seeking through GAT-3 upregulation and morphological plasticity at D1-MSN terminals. Mol Psychiatry. 2022;27(2):855–64.

114. Kruyer A, Scofield MD, Wood D, Reissner KJ, Kalivas PW. Heroin Cue-Evoked Astrocytic Structural Plasticity at Nucleus Accumbens Synapses Inhibits Heroin Seeking. Biol Psychiatry. 2019;86(11):811–9.

115. Luscher C, Malenka RC. Drug-evoked synaptic plasticity in addiction: from molecular changes to circuit remodeling. Neuron. 2011;69(4):650–63.

116. Thomas MJ, Kalivas PW, Shaham Y. Neuroplasticity in the mesolimbic dopamine system and cocaine addiction. Br J Pharmacol. 2008;154(2):327–42.

117. Kalivas PW, Volkow N, Seamans J. Unmanageable motivation in addiction: a pathology in prefrontal-accumbens glutamate transmission. Neuron. 2005;45(5):647–50.

118. Palhano-Fontes F, Andrade KC, Tofoli LF, Santos AC, Crippa JA, Hallak JE, et al. The psychedelic state induced by ayahuasca modulates the activity and connectivity of the default mode network. PLoS One. 2015;10(2):e0118143.

119. Zhang R, Volkow ND. Brain default-mode network dysfunction in addiction. Neuroimage. 2019;200:313–31.

120. Liang X, He Y, Salmeron BJ, Gu H, Stein EA, Yang Y. Interactions between the salience and default-mode networks are disrupted in cocaine addiction. J Neurosci. 2015;35(21):8081–90.

121. Li Q, Li Z, Li W, Zhang Y, Wang Y, Zhu J, et al. Disrupted Default Mode Network and Basal Craving in Male Heroin-Dependent Individuals: A Resting-State fMRI Study. J Clin Psychiatry. 2016;77(10):e1211–e7.

122. Li W, Li Q, Wang D, Xiao W, Liu K, Shi L, et al. Dysfunctional Default Mode Network in Methadone Treated Patients Who Have a Higher Heroin Relapse Risk. Sci Rep. 2015;5:15181.

123. Sheline YI, Barch DM, Price JL, Rundle MM, Vaishnavi SN, Snyder AZ, et al. The default mode network and self-referential processes in depression. Proc Natl Acad Sci U S A. 2009;106(6):1942–7.

124. Lanius RA, Bluhm RL, Coupland NJ, Hegadoren KM, Rowe B, Theberge J, et al. Default mode network connectivity as a predictor of post-traumatic stress disorder symptom severity in acutely traumatized subjects. Acta Psychiatr Scand. 2010;121(1):33–40.

125. Carhart-Harris RL, Erritzoe D, Williams T, Stone JM, Reed LJ, Colasanti A, et al. Neural correlates of the psychedelic state as determined by fMRI studies with psilocybin. Proc Natl Acad Sci U S A. 2012;109(6):2138–43.

126. Siegel JS, Subramanian S, Perry D, Kay BP, Gordon EM, Laumann TO, et al. Psilocybin desynchronizes the human brain. Nature. 2024.

127. Mertens LJ, Wall MB, Roseman L, Demetriou L, Nutt DJ, Carhart-Harris RL. Therapeutic mechanisms of psilocybin: Changes in amygdala and prefrontal functional connectivity during emotional processing after psilocybin for treatment-resistant depression. J Psychopharmacol. 2020;34(2):167–80.

128. Roseman L, Demetriou L, Wall MB, Nutt DJ, Carhart-Harris RL. Increased amygdala responses to emotional faces after psilocybin for treatment-resistant depression. Neuropharmacology. 2018;142:263–9.

129. Kraehenmann R, Preller KH, Scheidegger M, Pokorny T, Bosch OG, Seifritz E, et al. Psilocybin-Induced Decrease in Amygdala Reactivity Correlates with Enhanced Positive Mood in Healthy Volunteers. Biol Psychiatry. 2015;78(8):572–81.

130. Urban MM, Stingl MR, Meinhardt MW. Mini-review: The neurobiology of treating substance use disorders with classical psychedelics. Front Neurosci. 2023;17:1156319.

131. Nichols DE. Hallucinogens. Pharmacol Ther. 2004;101(2):131–81.

132. Nichols DE. Psychedelics. Pharmacol Rev. 2016;68(2):264–355.

133. Vollenweider FX, Vollenweider-Scherpenhuyzen MF, Babler A, Vogel H, Hell D. Psilocybin induces schizophrenia-like psychosis in humans via a serotonin-2 agonist action. Neuroreport. 1998;9(17):3897–902.

134. Sithurandi U, Wijayaratna D, Kankanamge D, Karunarathne A. Chapter Two - Molecular regulation of PLCβ signaling. In: Shukla A, editor. Methods in Enzymology. 682. Cambridge, MA: Academic Press; 2023. p. 17-52.

135. Yasuda R, Hayashi Y, Hell JW. CaMKII: a central molecular organizer of synaptic plasticity, learning and memory. Nat Rev Neurosci. 2022;23(11):666–82.

136. Saxena A, Scaini G, Bavaresco DV, Leite C, Valvassori SS, Carvalho AF, et al. Role of Protein Kinase C in Bipolar Disorder: A Review of the Current Literature. Mol Neuropsychiatry. 2017;3(2):108–24.

137. Fink CC, Meyer T. Molecular mechanisms of CaMKII activation in neuronal plasticity. Curr Opin Neurobiol. 2002;12(3):293–9.

138. Vargas MV, Dunlap LE, Dong C, Carter SJ, Tombari RJ, Jami SA, et al. Psychedelics promote neuroplasticity through the activation of intracellular 5-HT2A receptors. Science. 2023;379(6633):700-6.

139. Moliner R, Girych M, Brunello CA, Kovaleva V, Biojone C, Enkavi G, et al. Psychedelics promote plasticity by directly binding to BDNF receptor TrkB. Nat Neurosci. 2023;26(6):1032–41.

140. Sanchez-Alegria K, Flores-Leon M, Avila-Munoz E, Rodriguez-Corona N, Arias C. PI3K Signaling in Neurons: A Central Node for the Control of Multiple Functions. Int J Mol Sci. 2018;19(12).

141. Wu YT, Tan HL, Huang Q, Ong CN, Shen HM. Activation of the PI3K-Akt-mTOR signaling pathway promotes necrotic cell death via suppression of autophagy. Autophagy. 2009;5(6):824–34.

142. Trollope AF, Gutierrez-Mecinas M, Mifsud KR, Collins A, Saunderson EA, Reul JM. Stress, epigenetic control of gene expression and memory formation. Exp Neurol. 2012;233(1):3–11.

143. Kowianski P, Lietzau G, Czuba E, Waskow M, Steliga A, Morys J. BDNF: A Key Factor with Multipotent Impact on Brain Signaling and Synaptic Plasticity. Cell Mol Neurobiol. 2018;38(3):579–93.

144. Olson DE. Biochemical Mechanisms Underlying Psychedelic-Induced Neuroplasticity. Biochemistry. 2022;61(3):127–36.

145. Calder AE, Hasler G. Towards an understanding of psychedelic-induced neuroplasticity. Neuropsychopharmacology. 2023;48(1):104–12.

146. Volkow ND, Tomasi D, Wang GJ, Logan J, Alexoff DL, Jayne M, et al. Stimulant-induced dopamine increases are markedly blunted in active cocaine abusers. Mol Psychiatry. 2014;19(9):1037–43.

147. Volkow ND, Wang GJ, Fowler JS, Logan J, Gatley SJ, Gifford A, et al. Prediction of reinforcing responses to psychostimulants in humans by brain dopamine D2 receptor levels. Am J Psychiatry. 1999;156(9):1440–3.

148. Volkow ND, Wang GJ, Fowler JS, Logan J, Gatley SJ, Hitzemann R, et al. Decreased striatal dopaminergic responsiveness in detoxified cocaine-dependent subjects. Nature. 1997;386(6627):830-3.

149. Nutt DJ, Lingford-Hughes A, Erritzoe D, Stokes PR. The dopamine theory of addiction: 40 years of highs and lows. Nat Rev Neurosci. 2015;16(5):305–12.

150. Martinez D, Carpenter KM, Liu F, Slifstein M, Broft A, Friedman AC, et al. Imaging dopamine transmission in cocaine dependence: link between neurochemistry and response to treatment. Am J Psychiatry. 2011;168(6):634–41.

151. Vollenweider FX, Preller KH. Psychedelic drugs: neurobiology and potential for treatment of psychiatric disorders. Nat Rev Neurosci. 2020;21(11):611–24.

152. Amato D, Canneva F, Cumming P, Maschauer S, Groos D, Dahlmanns JK, et al. A dopaminergic mechanism of antipsychotic drug efficacy, failure, and failure reversal: the role of the dopamine transporter. Molecular Psychiatry. 2020;25(9):2101–18.

153. Amato D, Kruyer A, Samaha AN, Heinz A. Hypofunctional Dopamine Uptake and Antipsychotic Treatment-Resistant Schizophrenia. Front Psychiatry. 2019;10:314.

154. Amato D, Natesan S, Yavich L, Kapur S, Muller CP. Dynamic regulation of dopamine and serotonin responses to salient stimuli during chronic haloperidol treatment. Int J Neuropsychopharmacol. 2011;14(10):1327–39.

155. Amato D, Vernon AC, Papaleo F. Dopamine, the antipsychotic molecule: A perspective on mechanisms underlying antipsychotic response variability. Neurosci Biobehav Rev. 2018;85:146–59.

156. Bass CE, Grinevich VP, Gioia D, Day-Brown JD, Bonin KD, Stuber GD, et al. Optogenetic stimulation of VTA dopamine neurons reveals that tonic but not phasic patterns of dopamine transmission reduce ethanol self-administration. Front Behav Neurosci. 2013;7:173.

157. NIDA. What are treatments for tobacco dependence? 2020 [Available from: https://nida.nih.gov/publications/research-reports/tobacco-nicotine-e-cigarettes/what-are-treatments-tobacco-dependence#:~:text=There%20are%20effective%20treatments%20that%20support%20tobacco%20cessation%2C,replacement%20therapy%20as%20well%20as%20bupropion%20and%20varenicline.

158. Di Ciano P, Guranda M, Lagzdins D, Tyndale RF, Gamaleddin I, Selby P, et al. Varenicline-Induced Elevation of Dopamine in Smokers: A Preliminary [(11)C]-(+)-PHNO PET Study. Neuropsychopharmacology. 2016;41(6):1513–20.

159. Stahl SM, Pradko JF, Haight BR, Modell JG, Rockett CB, Learned-Coughlin S. A Review of the Neuropharmacology of Bupropion, a Dual Norepinephrine and Dopamine Reuptake Inhibitor. Prim Care Companion J Clin Psychiatry. 2004;6(4):159–66.

160. Zakiniaeiz Y, Liu H, Gao H, Najafzadeh S, Ropchan J, Nabulsi N, et al. Nicotine Patch Alters Patterns of Cigarette Smoking-Induced Dopamine Release: Patterns Relate to Biomarkers Associated With Treatment Response. Nicotine Tob Res. 2022;24(10):1597–606.

161. Sakashita Y, Abe K, Katagiri N, Kambe T, Saitoh T, Utsunomiya I, et al. Effect of psilocin on extracellular dopamine and serotonin levels in the mesoaccumbens and mesocortical pathway in awake rats. Biol Pharm Bull. 2015;38(1):134–8.

162. Wojtas A, Bysiek A, Wawrzczak-Bargiela A, Mackowiak M, Golembiowska K. Limbic System Response to Psilocybin and Ketamine Administration in Rats: A Neurochemical and Behavioral Study. Int J Mol Sci. 2023;25(1).

163. O’Shea E, Escobedo I, Orio L, Sanchez V, Navarro M, Green AR, et al. Elevation of ambient room temperature has differential effects on MDMA-induced 5-HT and dopamine release in striatum and nucleus accumbens of rats. Neuropsychopharmacology. 2005;30(7):1312–23.

164. Piras G, Cadoni C, Caria F, Pintori N, Spano E, Vanejevs M, et al. Characterization of the Neurochemical and Behavioral Effects of the Phenethylamine 2-Cl-4,5-MDMA in Adolescent and Adult Male Rats. Int J Neuropsychopharmacol. 2024;27(5).

165. Pettit HO, Justice JB, Jr. Dopamine in the nucleus accumbens during cocaine self-administration as studied by in vivo microdialysis. Pharmacol Biochem Behav. 1989;34(4):899–904.

166. Kankaanpaa A, Meririnne E, Lillsunde P, Seppala T. The acute effects of amphetamine derivatives on extracellular serotonin and dopamine levels in rat nucleus accumbens. Pharmacol Biochem Behav. 1998;59(4):1003–9.

167. Narita M, Aoki K, Takagi M, Yajima Y, Suzuki T. Implication of brain-derived neurotrophic factor in the release of dopamine and dopamine-related behaviors induced by methamphetamine. Neuroscience. 2003;119(3):767–75.

168. O’Brien CP. Drug Use Disorders and Addiction. In: Brunton LL, Hilal-Dandan R, Knollmann BC, editors. Goodman & Gilman’s: The Pharmacological Basis of Therapeutics, 13e. New York, NY: McGraw-Hill Education; 2017.

169. Schultz W. Getting formal with dopamine and reward. Neuron. 2002;36(2):241–63.

170. Volkow ND, Wang GJ, Fowler JS, Logan J, Gatley SJ, Wong C, et al. Reinforcing effects of psychostimulants in humans are associated with increases in brain dopamine and occupancy of D(2) receptors. J Pharmacol Exp Ther. 1999;291(1):409–15.

171. Wanat MJ, Willuhn I, Clark JJ, Phillips PE. Phasic dopamine release in appetitive behaviors and drug addiction. Curr Drug Abuse Rev. 2009;2(2):195–213.

172. Bungay PM, Morrison PF, Dedrick RL. Steady-state theory for quantitative microdialysis of solutes and water in vivo and in vitro. Life Sci. 1990;46(2):105–19.

