## Supplemental Data for "Psychedelics as pharmacotherapeutics for substance use disorders: a scoping review on clinical trials and perspectives on underlying neurobiology"

**Scoping Review Methodology:**

We conducted two systematic searches with the goals of characterizing ongoing, registered clinical trials that utilize a classic psychedelic intervention for treating SUDs and to identify the accumbal dopaminergic consequence following a systemic administration of a classic psychedelic in rodents. These searches are independently discussed as “Scoping Review of Ongoing Clinical Trials” and “Scoping Review of Microdialysis Data” respectively below. Both systematic literature searches were not registered or funded. For microdialysis data, studies included received funding from National Science Centre (2020/37/B/NZ7/03753) and Maj Institute of Pharmacology (1); Ministry of Science and Technology (SAF2001-1437) (2) and Ministry of Health (Grant FIS02/1185, G03/005).

*Scoping Review of Ongoing Clinical Trials:*

The goal of this scoping review was to identify all ongoing, registered clinical trials at clinicaltrials.gov that were utilizing a psychedelic intervention and measuring an outcome related to SUD (like Timeline Follow Back) to better understand the current state of clinical research in the field. Characterization of these trials can help inform clinicians and researchers to aid in predicting the outlook of research in the field, identify understudied outcomes and demographics, and develop new clinical trials with protocols based on ongoing clinical trials. For this search, we searched clinicaltrials.gov with combinations of the following criteria:

- Condition/Disease: Substance Use Disorders; Addiction
- Other Terms: None entered
- Intervention/Treatment: Psilocybin; DMT; LSD; Mescaline; Ibogaine; MDMA; Psychedelic

Because we were interested in all types of experimental design and application of psychedelic interventions, expert searches were not conducted to avoid not capturing all clinical trials. Further, we included all years, languages, trial statuses, ages, sexes, and those including health volunteers in our initial searches. After conducting grey searches for individual subtypes of SUD (ex: cocaine use disorder) and other terms related to specific drug use (ex: alcoholism) with each intervention included, two additional clinical trials were found (NCT06102434 and NCT03380728) utilizing psilocybin intervention for cocaine use disorder and ibogaine intervention for alcoholism, respectively. All searches occurred in August 2024.

<u>Example of a search used:</u>

Condition/Disease: Substance Use Disorders
Other Terms: None entered
Intervention/Treatment: Psychedelic
Set “First Posted” to August 1, 2024 to reproduce search results
Number of Hits: 53
Data Extraction: NCT Number, Study URL

Once all clinical trials were identified, the following exclusion criteria were applied: duplicate trial, lacking psychedelic intervention, not using an included drug such as cannabis, and not measuring drug use (Supp. Figure 1). Duplicates were identified by repeating NCT number across searches and study criteria including Trial ID, Sponsor, Phase, Status, Psychedelic Used, Dose Type and Frequency, Objective, Endpoint, Outcomes, Sample Size, Experimental Design, Sex Included, and Reported Expected Study Completion Date were manually identified to ensure proper characterization. Results were charted by L.W. and verified by all co-authors and readers.

***Supp Figure 1: Sorting Workflow for Clinical Trial Search Results***

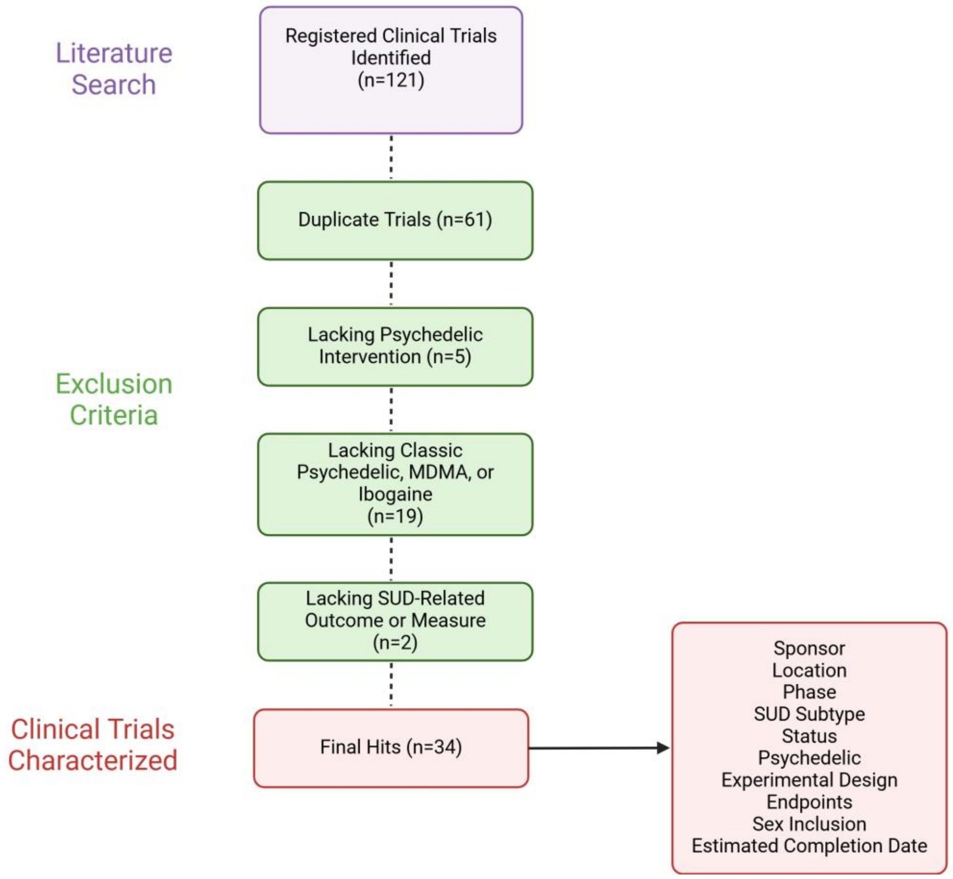

*Scoping Review of Microdialysis Data:*

Because the initial stages of SUD development is dependent on dopaminergic changes within the nucleus accumbens core associated with drug use, we sought to understand the dopaminergic consequence of psychedelic administration. To do accomplish this, we conducted systematic searches using PubMED and SCOPUS databases to identify published manuscripts using *in vivo* microdialysis to detect psychedelic-induced dopaminergic changes in the nucleus accumbens of animals. Grey literature searches were conducted on Google Scholar to identify any possible relevant publications not captured by our searches or registered in PubMed or SCOPUS. We took the first 20 hits from each grey literature search, added the term "striatum", and took any unique publications within the first 20 hits of the specified search. All years, language, and publication status was included in our initial searches to ensure we captured all publications that met stringent inclusion criteria. The last search conducted occurred in August 2024, so any attempts to reproduce our searches should use 9/1/2024 as their latest upload date when possible.

<u>Example of a search used (PubMed):</u>

Advanced Query:

("hallucinogens"[Mesh] OR "hallucinogen*" OR "psychedelic*" OR "psychotomimetic*") AND ("microdialysis"[Mesh] OR "microdialysis" OR "micro-dialysis" OR "micro dialysis") AND ("dopamine"[Mesh] OR "dopamin*" OR "3,4-Dihydroxyphenethylamine" OR "3,4 Dihydroxyphenethylamine") AND ("neostriatum"[Mesh] OR "corpus striatum"[Mesh] OR "neostriatum" OR "striatum" OR "accumbens" OR "cortex") AND ("Rodentia"[Mesh] OR "rodent*" OR "beaver*" OR "capybara*" OR "jerboa*" OR "mouse" OR "mice" OR "rat" OR "rats" OR "hydrochaeri*")

Number of hits: 68

After all publications were identified, all publications that lacked *in vivo* microdialysis data, relevance for the goal of the search, or duplicates were excluded (Supp. Fig. 2). More stringent exclusion criteria were then applied for studies that did not measure dopamine changes, were outside of the nucleus accumbens, or used multiple administrations of a psychedelic within one session. Due to a greater number of studies using rats and reporting dopamine differences as percentage change compared to baseline, we then excluded studies using mice to avoid species differences in the limited number of studies, and other units of measurement to avoid inappropriate extrapolation of data. Of the publications left, we then excluded any experiments that did not use a classic psychedelic or MDMA. Three final hits were detected that used psilocin, psilocybin, and MDMA and are described in Supp. Table 1. Data (the reported changes in dopamine induced by a psychedelic or vehicle control) was then replotted using WebPlot Digitizer, extracted into an Excel document, and then graphed as shown in Figure 3. Due to differences in the length of experiment across studies, only the first 3 hours following drug administration is reported. We included one additional study that used an MDMA derivative (3) in our data extraction, however only data from their vehicle control group was used to balance our vehicle group between psilocin/psilocybin and MDMA publications (Supp. Table 2). Results were charted by L.W. and verified by all co-authors and readers.

***Supp Figure 2: Sorting Workflow for In Vivo-Microdialysis Search Results***

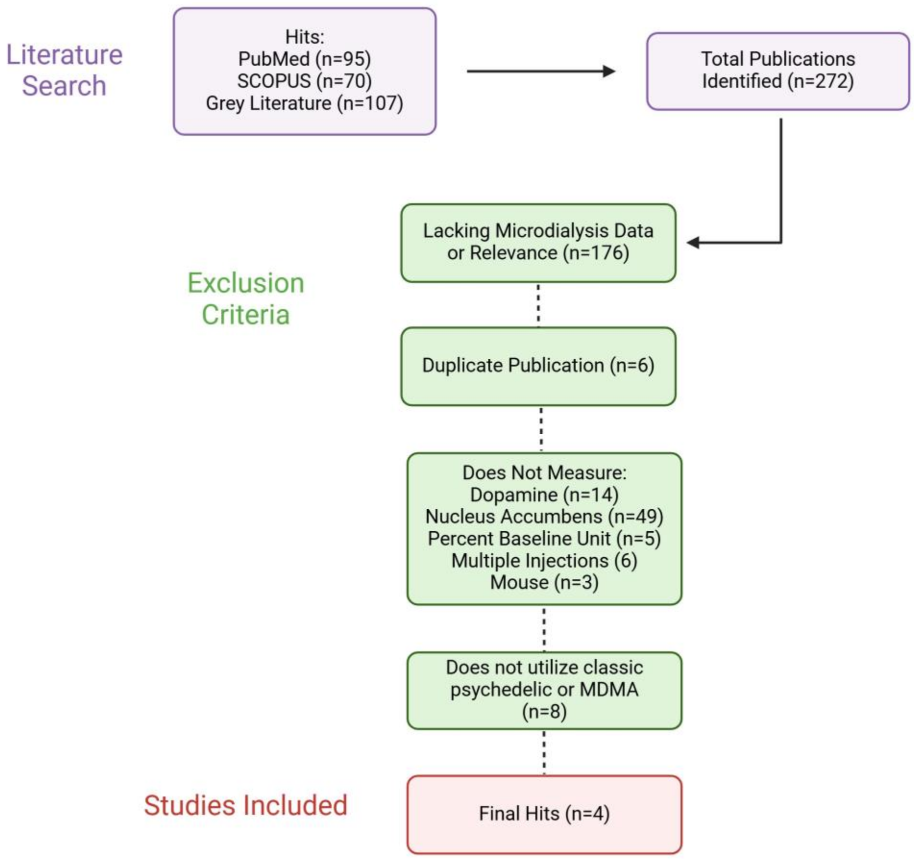

**Supplementary Table 1: Positive Hits from "Scoping Review of Microdialysis Data"**

| Authors | Species (Strain) | Drug(s) Used | Dose | Groups extracted |
|---|---|---|---|---|
| Sakashita et al. (4) | Rat (Wistar) | Psilocin | 1, 5, 10 mg/kg i.p. | Vehicle; 1, 5, 10 mg/kg psilocin |
| Wojitas et al. (1) | Rat (Wistar-Han) | Psilocybin<br>Ketamine | 2, 10 mg/kg i.p.<br>10mg/kg i.p. | Vehicle, 10mg/kg psilocybin |
| O'Shea et al. (2) | Rat (Dark Agouti) | MDMA | 2.5, 5mg/kg i.p. | Vehicle; 2.5, 5mg/kg (20C) |
| Piras et al. (3) | Rat (Sprague-Dawley) | 2-Cl-4,5-MDMA | 1, 3, 5mg/kg i.v. | Vehicle (Adults) |

**Supplementary Table 2: Data Extracted From Positive Hits**

| Authors | Group | -40 | -20 | 0 | 20 | 40 | 60 | 80 | 100 | 120 | 140 | 160 | 180 |
|---|---|---|---|---|---|---|---|---|---|---|---|---|---|
| Sakashita et al. (4) | Vehicle | 91.361 | 97.278 | 107.93 | 97.278 | 94.32 | 85.444 | 102.6 | 93.136 | 98.462 | 91.953 | 100.83 | 99.053 |
| | 1mg/kg psilocin | 93.136 | 103.79 | 102.6 | 93.136 | 92.544 | 94.32 | 102.6 | 96.686 | 93.728 | 103.79 | 94.32 | 93.136 |
| | 5mg/kg psilocin | 102.01 | 96.686 | 100.83 | 116.8 | 106.15 | 97.87 | 90.769 | 87.811 | 86.627 | 91.361 | 88.402 | 86.627 |
| | 10mg/kg psilocin | 102.01 | 103.79 | 97.278 | 110.89 | 139.29 | 138.11 | 123.91 | 112.66 | 112.66 | 108.52 | 98.462 | 103.2 |
| Wojitas et al. (1) | Vehicle | 99.097 | 99.186 | 100.56 | 105.79 | 104.59 | 98.24 | 99.616 | 100.98 | 103.65 | 102.44 | 101.25 | 102.62 |
| | 10mg/kg psilocybin | 109.4 | 99.186 | 100.56 | 168.88 | 134.2 | 140.73 | 149.83 | 137.04 | 162.87 | 177.13 | 178.5 | 164.43 |
| O'Shea et al. (2) | Vehicle for 2.5 (20C) | 105.09 | 99.99 | 97.52 | 95.05 | 89.94 | 84.86 | 74.52 | 72.05 | 72.19 | 74.96 | 80.35 | 83.1 |
| | 2.5mg/kg MDMA (20C) | 102.47 | 99.99 | 97.52 | 97.66 | 97.81 | 100.58 | 98.1 | 100.87 | 101.01 | 101.16 | 101.3 | 101.45 |
| | Vehicle for 5 (20C) | 82.19 | 87.67 | 65.75 | 84.93 | 98.63 | 71.23 | 95.89 | 106.8 | 104.1 | 104.1 | 71.23 | 65.75 |
| | 5mg/kg MDMA (20C) | 106.8 | 95.89 | 84.93 | 197.3 | 265.8 | 375.3 | 298.6 | 227.4 | 158.9 | 147.9 | 137 | 131.5 |
| Piras et al. (3) | Vehicle | N/A | N/A | 99.335 | 94.366 | 99.498 | 88.756 | 92.443 | 109.83 | 101.98 | 91.958 | 86.27 | 98.611 |

### References:


1. Wojtas A, Bysiek A, Wawrzczak-Bargiela A, Mackowiak M, Golembiowska K. Limbic System Response to Psilocybin and Ketamine Administration in Rats: A Neurochemical and Behavioral Study. Int J Mol Sci. 2023;25(1).
2. O'Shea E, Escobedo I, Orio L, Sanchez V, Navarro M, Green AR, et al. Elevation of ambient room temperature has differential effects on MDMA-induced 5-HT and dopamine release in striatum and nucleus accumbens of rats. Neuropsychopharmacology. 2005;30(7):1312-23.
3. Piras G, Cadoni C, Caria F, Pintori N, Spano E, Vanejevs M, et al. Characterization of the Neurochemical and Behavioral Effects of the Phenethylamine 2-Cl-4,5-MDMA in Adolescent and Adult Male Rats. Int J Neuropsychopharmacol. 2024;27(5).
4. Sakashita Y, Abe K, Katagiri N, Kambe T, Saitoh T, Utsunomiya I, et al. Effect of psilocin on extracellular dopamine and serotonin levels in the mesoaccumbens and mesocortical pathway in awake rats. Biol Pharm Bull. 2015;38(1):134-8.
