## Supplementary figures and images for "Psychedelics as pharmacotherapeutics for substance use disorders: a scoping review on clinical trials and perspectives on underlying neurobiology"

### Supplemental Figure 1

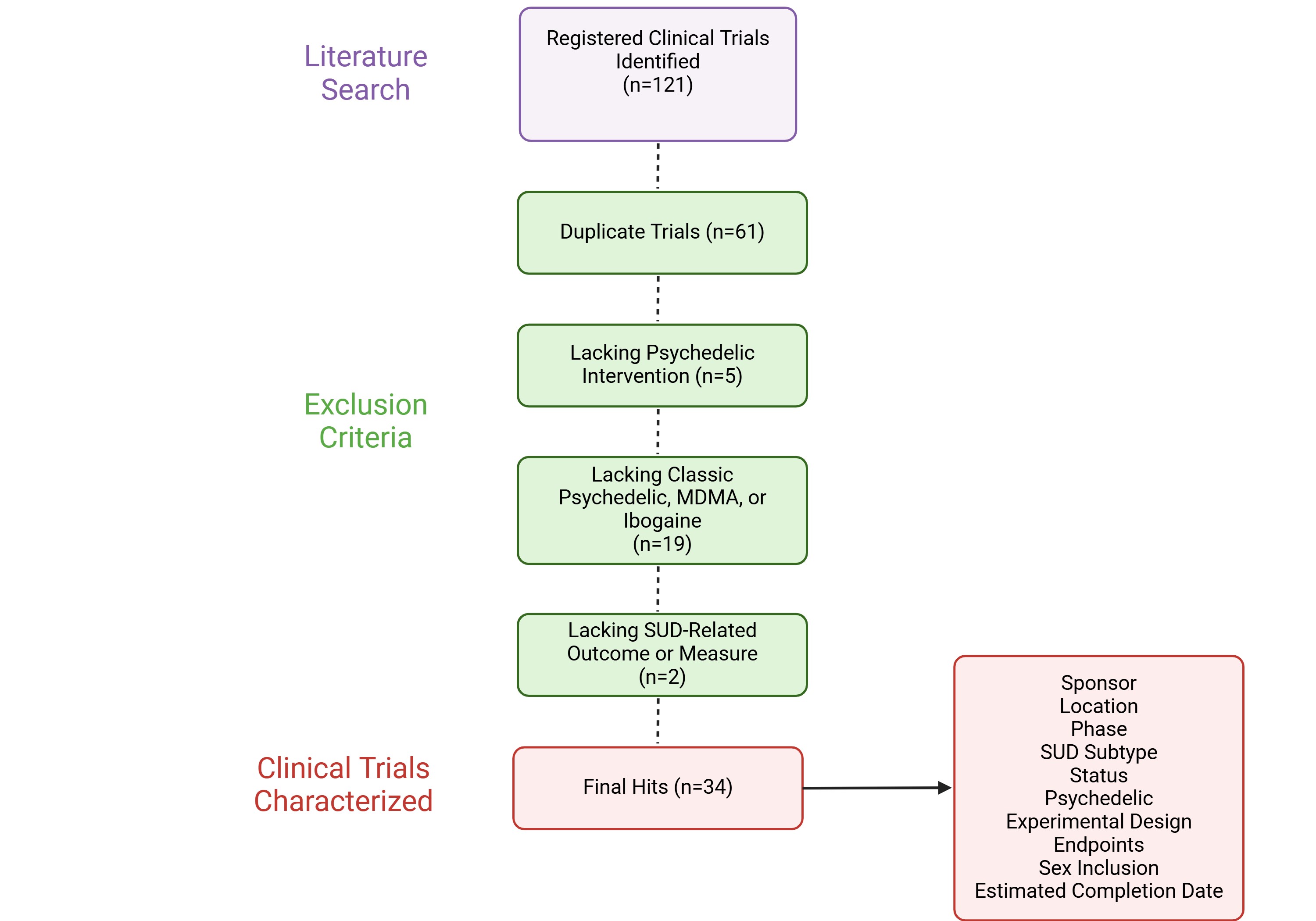

### Supplemental Figure 2

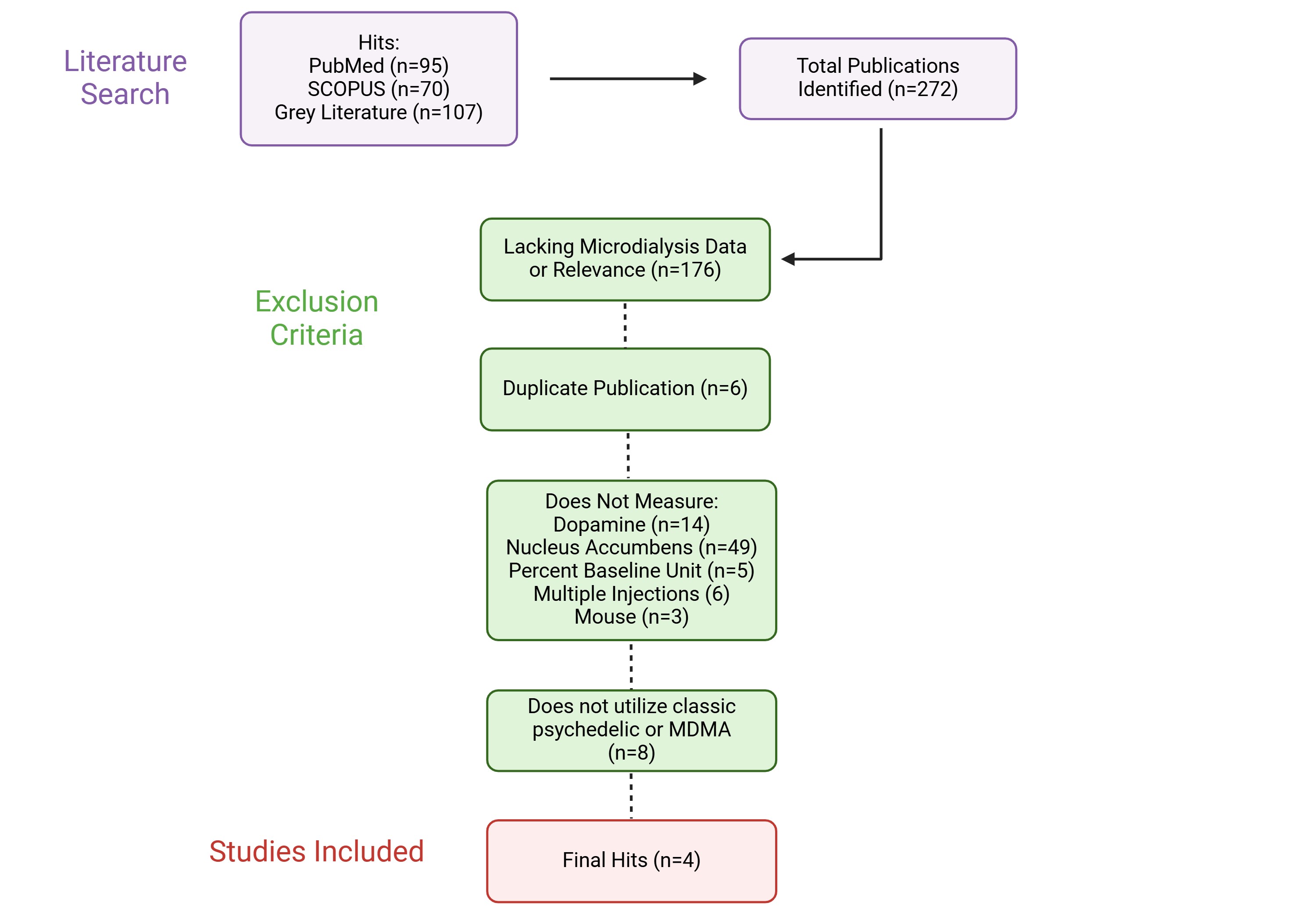
